# Performance boundaries of plasma-centred prediction across PET- and MRI-defined AT(N): QC-first internal validation in ADNI with separate axis-specific assessments

**DOI:** 10.64898/2026.10.15.26365631

**Authors:** Jiayuan Xu, Sofia Michopoulou, Fumie Costen, the Alzheimer’s Disease Neuroimaging Initiative

## Abstract

**Background:** Plasma p-tau217 identifies Alzheimer’s disease pathology, but performance in mixed cohorts may not extend to conditional tau-positivity discrimination within amyloid-positive individuals or to MRI-defined hippocampal neurodegeneration. We compared plasma prediction across PET- and MRI-defined AT(N) outcomes and conducted separate axis-specific assessments.

**Methods:** This retrospective secondary study used Alzheimer’s Disease Neuroimaging Initiative (ADNI) data. Tasks were amyloid positivity (A; 1,097 participants, 496 positive), tau positivity (T; 929, 195), T within A+ (361, 161), study-defined hippocampal neurodegeneration (N; 1,303, 248), and joint A+T+ (852, 161). Repeated nested cross-validation used fold-internal preprocessing and calibration. Post-hoc assessments comprised a Bio-Hermes A-axis staged random-forest analysis using Lilly p-tau217 and C2N A*`β`*40/A*`β`*42 (908 participants; 328 and 324 positive under the primary and alternative PET classifications), and an A4 ordinal p-tau217 analysis against an A4-specific medial-temporal tau-PET endpoint in A+ participants.

**Results:** ADNI random-forest AUROCs were 0.9425 (95% CI 0.9271–0.9562) for A, 0.9495 (0.9325–0.9645) for T, 0.8852 (0.8485–0.9163) for T within A+, 0.7865 (0.7553–0.8151) for N, and 0.9614 (0.9484–0.9726) for joint A+T+. At thresholds targeting 90% sensitivity, specificity was 87.5% for T overall and 60.7% within A+. In Bio-Hermes, adding A*`β`*40 and A*`β`*42 to p-tau217 increased AUROC from 0.894 to 0.916 (paired difference 0.022, 95% conditional interval 0.010– 0.035) and changed Brier score by **-0.011** (**-0.017** to **-0.005**); the alternative PET classification was concordant. In A4, ordinal p-tau217 had AUROC 0.7356 (95% CI 0.6775–0.7907), average precision 0.5657 (0.4732–0.6677), and Spearman ***ρ* = 0.4718** (0.3824–0.5528).

**Conclusions:** ADNI identified task-dependent performance boundaries, particularly attenuated conditional T discrimination within A+. Separate-cohort evidence supported moderate single-marker conditional-T discrimination in A4 and modest complementary A-axis information from plasma A*`β`* in Bio-Hermes, but only within those respective axes and assay implementations.

## 1 Background

The amyloid/tau/neurodegeneration framework distinguishes amyloid deposition, pathological tau, and neurodegeneration from the clinical syndrome [1, 2]. The 2024 revised criteria further distinguish early-changing Core 1 markers, including accurate plasma p-tau217 tests, from later-changing Core 2 markers that more directly inform biological stage; structural MRI measures of neurodegeneration remain downstream and nonspecific [3]. A blood marker that identifies broad Alzheimer’s disease pathology may therefore perform differently when asked to discriminate tau positivity among people who are already amyloid positive or to classify MRI-defined neurodegeneration.

Plasma p-tau217 discriminates amyloid and overall tau-PET positivity well across research cohorts [4, 5]. The distinction between broad pathology detection and finer-grained conditional classification is less settled. Published studies report lower performance after restriction to amyloid-positive participants and weaker separation of adjacent PET-defined stages than of broadly separated groups [6–8]. Studies of automated plasma p-tau217 assays and A/T profiles have also shown that strong binary pathology detection does not imply precise regional or multilevel PET staging [9, 10]. This pattern is biologically plausible because plasma p-tau217 rises early with amyloid-related disease processes, whereas later-changing tau species such as plasma MTBR-tau243 more directly track insoluble tau aggregates [3, 11]. Neurodegeneration poses a different problem because hippocampal atrophy can reflect age, comorbidity, premorbid brain structure or reserve, and non-Alzheimer processes.

Our group previously used plasma p-tau181, neurofilament light, amyloid measures, APOE, cognition, and machine learning to predict amyloid-PET positivity, with an external dataset used for evaluation [12]. In contrast, the present study centres on p-tau217 and compares A, T, a study-defined hippocampal N measure, T within A+, joint A+T+, and A/T profiles under one QC-first repeated nested internal-validation framework, using predictors that exclude cognition, diagnosis, and cerebrospinal-fluid biomarkers. It aims to characterise pathology aligned with the AT(N) framework rather than the clinical syndrome of Alzheimer’s disease.

Flexible multivariable models can appear optimistic if tuning and evaluation use the same observations; nested cross-validation separates model selection from heldout performance estimation [13]. Even with leakage control, a high AUROC does not establish transportable probabilities, validated thresholds, or clinical utility [14, 15]. The novelty claimed here is not the first plasma prediction of A/T profiles or the first report of weaker discrimination within A+. It is the unified QC-first comparison of A, overall T, conditional T within A+, study-defined hippocampal N, joint A+T+, predictor blocks, and endpoint-definition sensitivity under one repeated nested internal-validation design. We treated the cross-axis comparison as exploratory benchmarking. We additionally conducted two post-hoc axis-specific assessments in cohorts separate from ADNI: a Bio-Hermes A-axis predictor-block assessment under different p-tau217 and A*β* assay implementations and PET classifications, and an A4 ordinal p-tau217 assessment of conditional tau positivity within A+ [16–18].

## 2 Methods

### 2.1 Study design and reporting

This retrospective secondary study used controlled-access ADNI data for model development and repeated internal validation. The fixed ADNI data cut was dated 21 June 2026. This retrospective secondary analysis was not prospectively registered. Reporting was prepared with reference to TRIPOD+AI, STARD 2015, and STROBE [19–21]. The cross-axis AT(N) comparison was exploratory and was not treated as a prespecified set of co-primary outcomes. The present purpose was research benchmarking and internal performance estimation; no deployable individual-level clinical decision tool or current care-pathway role was proposed.

After preliminary inspection of A-axis results in the same cohort, random forest (RF) was selected as the A-axis reporting model. We subsequently used RF as a common descriptive reporting model across axes while retaining the complete model–task grid. RF-versus-logistic contrasts compare complete predictor-and-model packages and therefore do not isolate the contribution of one biomarker. Because model-family selection used the same cohort, the RF results are post-selection rather than fully prespecified.

### 2.2 ADNI participants and plasma anchor

ADNI is a longitudinal multicentre programme for development and standardisation of clinical, imaging, genetic, and biofluid markers in Alzheimer’s disease [22–24]. We constructed one record per participant. The plasma anchor was the earliest collection with complete Fujirebio measurements of p-tau217, A*β*42, and A*β*40 after the plausibility cleaning detailed in the Supplementary Methods (Additional file 1). Field-specific missing codes were treated as missing, while valid negative imaging values were retained. Analysis eligibility required measurements of p-tau217, A*β*42, A*β*40, age, sex, APOE *ε*4 allele count, and the relevant outcome. All eight predictors were complete in the final outcome-specific cohorts; fold-internal median imputation remained in every model pipeline to preserve leakage-safe processing if missingness arose.

Amyloid PET, flortaucipir tau PET, FreeSurfer-derived hippocampal MRI measures, cognitive measures, and diagnosis were aligned separately to the plasma date. Imaging candidates had to lie within *±*365 days. For both amyloid and tau PET, raw candidates were first restricted to University of California, Berkeley qc_flag=2 (pass) or 1 (partial pass with the primary region of interest usable); tau candidates were additionally restricted to flortaucipir. Failed, unassessed, unprocessable, unknown, and missing QC categories were excluded before temporal matching. The nearest eligible scan was then selected deterministically. The QC flag of the selected scan was retained as provenance metadata and excluded from every feature matrix. Release filenames, value-cleaning rules, tie handling, and modality-specific QC policies are detailed in the Supplementary Methods (Additional file 1).

### 2.3 Plasma measurements

According to the ADNI release documentation, EDTA plasma aliquots had been analysed once, without technical replicates, for p-tau217, A*β*42, and A*β*40 on the Fujirebio Lumipulse G1200 automated chemiluminescent enzyme-immunoassay platform. The release documentation records storage, thawing, QC, and reagent-lot procedures. The measurements spanned multiple analytical runs and reagent lots; model performance was not evaluated separately by lot.

### 2.4 Reference outcomes

The amyloid outcome (A) was the released AMYLOID_STATUS on the selected QC-eligible UCBERKELEY_AMY_6MM record. The released field applies tracer-specific cortical-summary standardized uptake value ratio (SUVR) thresholds within the harmonised PET pipeline described in the dataset methods documentation [25].

Tau positivity (T) was defined from flortaucipir temporal meta-region-of-interest SUVR. The threshold was the mean plus two standard deviations in 361 dual-PET-QC-filtered amyloid-negative cognitively normal reference participants (full-precision cutoff 1.3498184). Overall T evaluated broad tau-positivity discrimination across all eligible participants. T within A+ used the same threshold after restriction to amyloid-positive participants and represented conditional discrimination between A+T- and A+T+, not regional or multilevel tau staging.

The neurodegeneration outcome (N) was specific to study-defined hippocampal atrophy. After exclusion of MRI records explicitly marked OVERALLQC=Fail and application of anatomical plausibility bounds, summed bilateral FreeSurfer hippocampal volume was regressed on intracranial volume, age, and sex in 411 amyloid-negative cognitively normal reference participants. Residuals below -1283.6734 mm^3^ were classified N+. A hippocampal-volume-to-intracranial-volume threshold of 0.00426101 was used in a fixed-score relabeling diagnostic. Joint A+T+ contrasted participants positive for both PET outcomes with all other participants who had both outcomes. This joint task measured a broad pathological profile and was not interpreted as isolated tau staging. T and N reference thresholds were estimated before model cross-validation; their full estimation uncertainty was not propagated through model development.

### 2.5 Predictors and model packages

The full package contained eight predictors: natural-log p-tau217, A*β*42, A*β*40, A*β*42/A*β*40, age, sex, education, and APOE *ε*4 allele count. Imaging variables, outcome labels, diagnoses, cognition, and cerebrospinal-fluid biomarkers were forbidden predictors.

We evaluated elastic-net logistic regression, RF, gradient boosting, and a radialbasis-function support-vector machine. These models were selected to span regularised linear, tree-ensemble, and kernel-based nonlinear approaches. Two single-predictor L2-logistic benchmarks used either log(p-tau217) alone or log[p-tau217/A*β*42] alone (*C* = 1, balanced class weights). These were study-defined research benchmarks and did not represent commercial or regulatory-cleared algorithms. Balanced class weights were used where specified to address outcome imbalance; probability calibration was estimated separately without class weighting. Every model used a pipeline containing median imputation and standardisation; the preprocessing parameters were fitted only within the relevant training split. The exact executed estimators, grids, software versions, class weights, selection rules, calibration inputs, and seeds are given in Additional file 1 (Supplementary Methods and Table S9).

### 2.6 Nested cross-validation, calibration, and operating points

Internal performance used stratified five-fold outer cross-validation repeated three times with random seed 42. Hyperparameters were selected by stratified three-fold inner cross-validation restricted to each outer-training set. Within each outer-training set, a second three-fold inner cross-validation generated out-of-fold decision scores. These scores were used to fit an L2-logistic sigmoid calibrator (*C* = 1, no class weighting) and to select Youden and target-sensitivity or target-specificity thresholds. The tuned learner was then refitted on the complete outer-training set. Its calibrator and thresholds were applied once to the held-out outer fold.

Each participant received one held-out probability and decision per repeat. Participant-level probabilities were averaged across the three repeats for AUROC, average precision (AP), and Brier score. Threshold metrics pooled held-out decisions while bootstrap resampling kept a participant’s repeated decisions together. For A, the 90%-sensitivity operating point was compared with the 2025 Alzheimer’s Association clinical-practice threshold in the closest intended-use sense, while recognising that ADNI included cognitively unimpaired participants and was not a specialty-care implementation cohort. The same 90%-sensitivity/75%-specificity threshold was applied to T, T within A+, N, and joint A+T+ only as an exploratory common performance reference, not as a guideline-endorsed criterion for those outcomes [26, 27]. We distinguished point-estimate threshold crossing from precision-supported attainment based on the lower confidence limits.

### 2.7 Statistical analysis

AUROC percentile confidence intervals (CIs) used 2,000 participant bootstrap resamples conditional on the realised averaged out-of-fold predictions and labels. For the reporting RF, an unpenalised joint logistic calibration model estimated intercept and slope from the participant-level probabilities averaged across repeats; 2,000 participant bootstrap resamples provided conditional percentile intervals. The two named parsimonious benchmark models were fixed for the final QC-filtered analysis after exploratory work in the same cohort; their contrasts with the selected RF remained post-selection comparisons. AUROC differences used 2,000 class-stratified paired-participant bootstrap resamples and paired DeLong tests [28]. The bootstrap intervals were not adjusted for multiple comparisons. Holm correction controlled family-wise error across only the two named A-axis DeLong *P* values. These CIs quantify participant-sampling uncertainty conditional on the realised out-of-fold predictions and labels; they do not include uncertainty from model-family selection, reference-threshold estimation, data splitting, tuning, calibration, or full model redevelopment.

Because this retrospective secondary analysis used all participants meeting outcome-specific eligibility and data-availability requirements in a fixed data cut, no prospective sample-size calculation was performed. Outcome-specific participant and positive-event counts and bootstrap CIs characterise uncertainty around the available estimates. Outcome cohorts differed in outcome-specific data availability, prevalence, and biomarker spectrum. Comparisons across A, T, N, and joint tasks, including overall T versus T within A+, were therefore descriptive; no formal cross-axis superiority test was performed. The complete model–task grid, selected sensitivity analyses, and disclosure-safe supporting results are provided in Additional file 1.

Cumulative RF predictor-block analyses evaluated p-tau217 alone, addition of the three plasma A*β* variables, addition of age, sex, and education, and addition of APOE *ε*4. A separate exploratory RF ablation excluded all plasma A*β* variables. Sensitivity analyses examined log transformations, alternative imaging windows and plasma-anchor rules, cognitively impaired participants, and descriptive subgroups. Fixed-score relabeling held the original out-of-fold scores fixed and changed only the T or N labels; it assessed dependence on endpoint definition rather than performance after model redevelopment under the alternative definitions. Label-permutation negative controls reran the modelling pathway after shuffling outcome labels as a check for gross leakage.

Multiclass models considered the joint A/T profile. The three-class analysis omitted the sparse A-T+ profile and was used as the interpretable summary; it therefore does not represent all four A/T profiles. The sparse cell count is not reported to comply with controlled-data disclosure requirements. Four-class results were descriptive only.

### 2.8 Post-hoc Bio-Hermes A-axis assessment

We conducted a post-hoc A-axis predictor-block assessment in controlled-access Bio-Hermes-001 data, a cohort separate from ADNI [16]. The common analysis-complete cohort included 908 participants and required Screening blood measurements, age, sex, education, APOE *ε*4 allele count, and baseline IXICO PET classifications with scan quality control recorded as pass. The primary endpoint was the released IXICO florbetapir visual-read classification (`AMYCLASV`; 328 positive); the alternative released IXICO classification (`AMYCLAS`; 324 positive) was evaluated as an endpoint-definition sensitivity. Both labels were used unchanged in the same participants. In the parent study, PET scans were centrally read by trained IXICO experts following the manufacturer’s read process with access to SUVR values. Disclosure-safe characteristics and analytic completeness were summarised for this same cohort.

Bio-Hermes blood biomarkers were measured in plasma samples; the released predictors comprised Lilly Clinical Diagnostics Laboratory p-tau217 and C2N PrecivityAD A*β*40 and A*β*42 measurements. Values were analysed on their released scales without cross-assay conversion. Cumulative predictor stages comprised: (1) Lilly p-tau217; (2) addition of C2N A*β*40 and A*β*42; (3) addition of age, sex, and education; and (4) addition of APOE *ε*4 allele count. Assay-censored p-tau217 values were retained as ordered tied categories without numerical substitution, and the empirical order transformation was fitted within each training fold. A random forest with 500 trees and balanced-subsample class weights was tuned over minimum leaf sizes 2, 5, and 10 by three-fold inner cross-validation. Evaluation used five outer folds repeated three times, with identical outer participant splits across stages within each endpoint.

Bio-Hermes models were developed and evaluated entirely within Bio-Hermes using native assay scales and raw random-forest probabilities. AUROC, AP, and Brier score were calculated for each complete set of outer-fold predictions within a repeat and then averaged across repeats. Stage 2-minus-Stage 1 was specified within the locked post-hoc assessment before formal fitting. Paired 95% percentile intervals were obtained from 2,000 participant resamples of the fixed repeated-crossfit predictions; models were not refitted during bootstrap.

### 2.9 A4 separate-cohort conditional tau assessment

The post-hoc A4 assessment used controlled-access data after prior exploratory A4 results had been viewed. The protocol was locked before the first formal analysis run. The analysis was restricted to A4 participants with official amyloid-positive screening status, an eligible Visit 6 Lilly electrochemiluminescence p-tau217 measurement collected no later than the treatment-start calendar day, and a dated quantitative baseline flortaucipir tau-PET scan. Visit 6 blood was protocol-scheduled before dosing, but the release did not contain participant-level collection and infusion times, so individual pre-dose order could not be verified [17]. The parent A4 trial was registered as NCT02008357.

The index marker was represented as an ordinal, censor-aware score. Values reported as below the lower limit of quantification were tied below every quantified value, quantified values retained their numerical order and ties, and values above the upper limit of quantification would have been tied above every quantified value. No concentration was imputed for assay-censored observations, and the A4 Lilly scale was not converted to the ADNI Fujirebio scale. A cohort-wise z-score was not analysed because a positive affine transformation of a single ordinal score preserves AUROC, AP, and rank correlation and would not establish assay commutability.

The earliest dated, analysed baseline quantitative flortaucipir scan was selected before applying the blood–scan interval criterion. The endpoint combined bilateral entorhinal and parahippocampal PERSI SUVRs using published atlas-region volume weights:

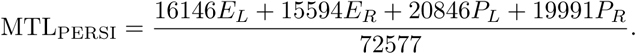

Here, *E_L_* and *E_R_* denote left and right entorhinal PERSI SUVRs, *P_L_* and *P_R_* denote left and right parahippocampal PERSI SUVRs, and the coefficients are the corresponding atlas-region volumes. A4-specific T positivity required MTL_PERSI_ *>* 1.03; equality remained negative. The threshold is an A4-derived research definition rather than an independent clinical standard [18]. The selected first baseline scan had to satisfy 0 *≤* blood day minus scan day *≤* 365; a later scan could not replace an earlier out-of-window scan.

The primary performance measure was AUROC for discrimination of A4-specific T status by ordinal p-tau217. Secondary summaries were AP, calculated with `average_precision_score`, and Spearman correlation with continuous MTL_PERSI_. Higher AP indicates better precision–recall performance, but AP depends on outcome prevalence and was not compared across tasks. Percentile intervals used 10,000 participant bootstrap resamples with a prespecified seed. Sensitivities restricted the imaging window to 180 days, retained quantified assay results only, or excluded sameday PET–blood pairs. A strict calendar-day pretreatment subset and exact-threshold equality were audited privately. The primary analysis required disclosure-safe support in both outcome classes; any sensitivity that could reveal a small cell protected by controlled-data disclosure rules, directly or by subtraction, was suppressed as a complete row. Because no ADNI probability model, assay scale, or operating threshold was transported to A4, the analysis did not estimate probability calibration, Brier score, clinical workpoints, PPV, NPV, application of an ADNI cutoff, or transported multivariable-model performance.

Additional file 1 provides the Supplementary Methods, aggregate ADNI cohort construction and selection (Tables S1–S2), complete binary performance (Tables S3– S4), staged predictor analyses (Table S5), operating-point and design sensitivities (Table S6), multiclass and negative controls (Table S7), fixed-score relabeling diagnostics (Table S8), executed model specifications (Table S9), the aggregate A4 assessment (Table S10), the post-hoc Bio-Hermes A-axis assessment (Table S11), ADNI RF calibration diagnostics (Table S12), and Bio-Hermes cohort characteristics and analytic completeness (Table S13), together with the ADNI precision–recall plot (Figure S1).

## 3 Results

### 3.1 Participants and outcome-specific cohorts

Among 4,728 raw ADNI amyloid-PET scans, 4,347 had QC flag 1 or 2 and 381 were excluded as failed, unassessed, unprocessable, unknown, or missing. QC-first nearest-date matching yielded 1,220 amyloid scans and 963 flortaucipir tau scans within *±*365 days of a plasma anchor. The anchored cohort comprised 1,592 participants (Table 1), of whom 1,522 had complete model predictors. Outcome-specific cohorts included 1,097 participants for A (496 positive), 929 for T (195 positive), 361 for T within A+ (161 positive), 1,303 for N (248 positive), and 852 for joint A+T+ (161 positive).

**Table 1.**
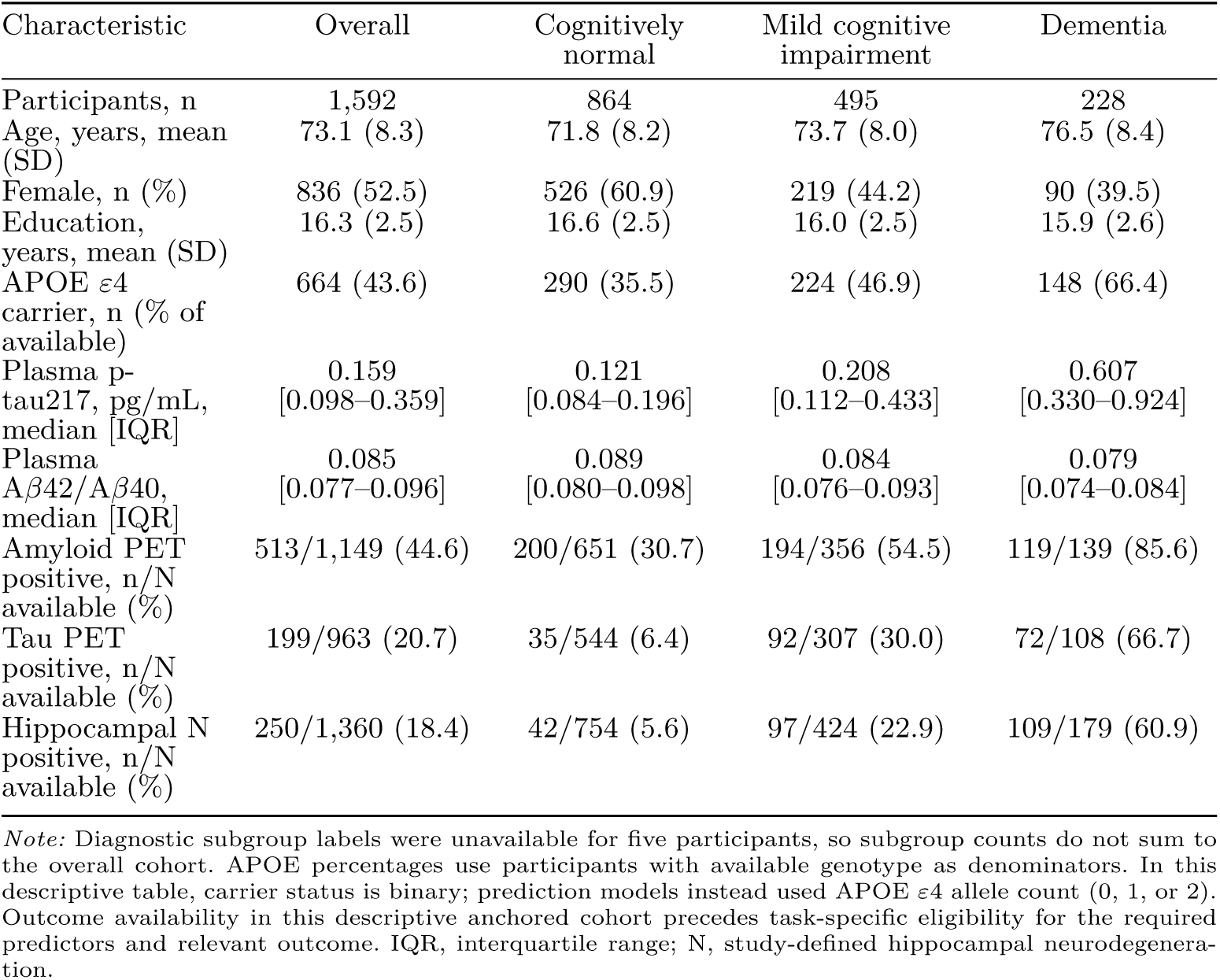
Characteristics of the plasma-anchored cohort.

### 3.2 Plasma prediction across AT(N) outcomes

The common RF package showed high internal discrimination for A, T overall, and joint A+T+, intermediate discrimination for T within A+, and lower discrimination for study-defined hippocampal N (Table 2; Figure 1). AUROC estimates were 0.9425 (95% conditional CI 0.9271–0.9562) for A, 0.9495 (0.9325–0.9645) for T, 0.8852 (0.8485–0.9163) for T within A+, 0.7865 (0.7553–0.8151) for N, and 0.9614 (0.9484–0.9726) for joint A+T+. These cross-axis contrasts are descriptive rather than paired superiority comparisons because the eligible samples, outcome prevalences, and biomarker spectra differed across prediction tasks.

**Fig. 1.**
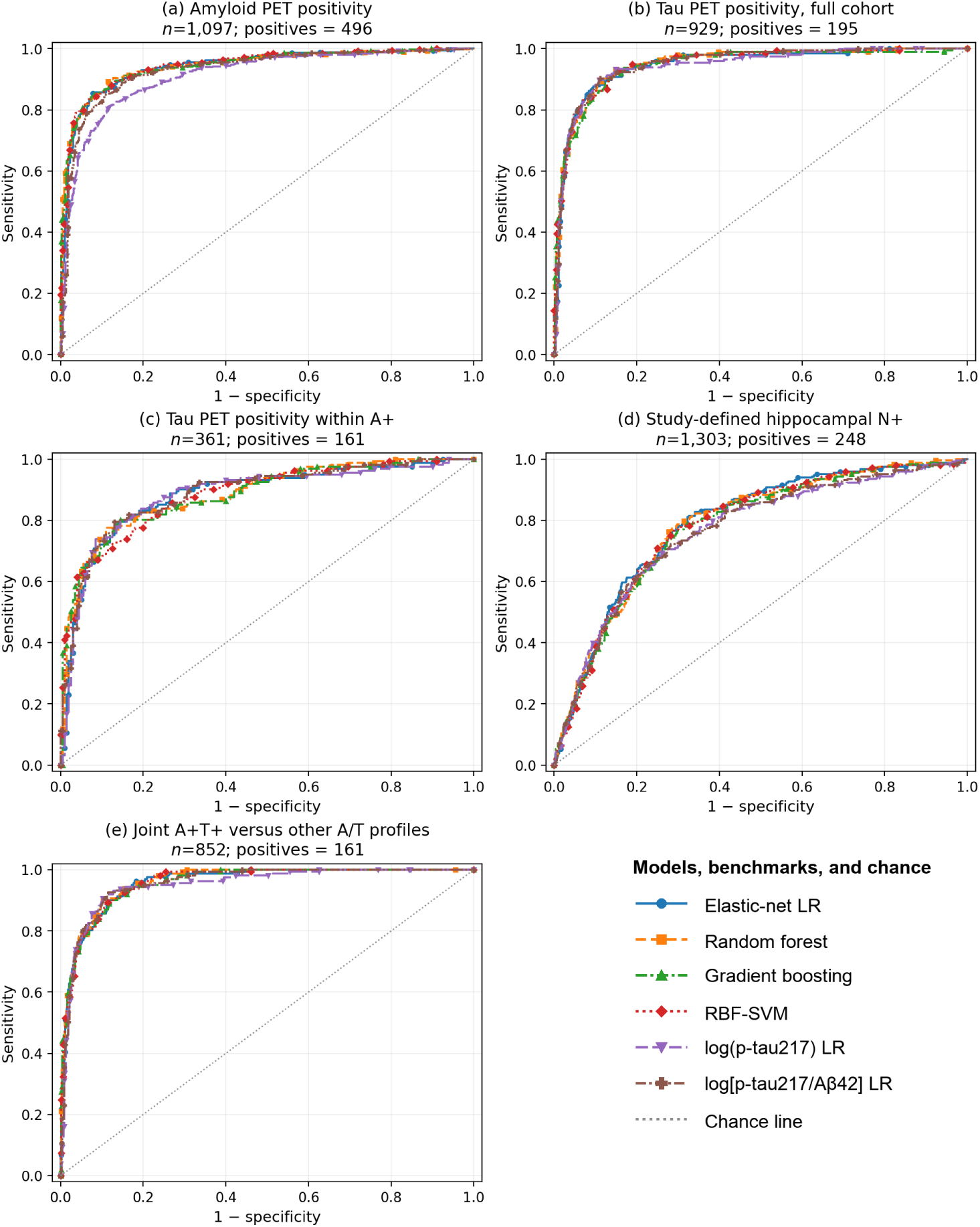
Internal-validation receiver-operating-characteristic curves across AT(N) outcomes. Panels show (a) amyloid-PET positivity, (b) tau-PET positivity in the full cohort, (c) tau-PET positivity among amyloid-positive participants, (d) study-defined hippocampal neurodegeneration, and (e) joint A+T+ versus other complete A/T profiles. Curves use participant-level out-of-fold probabilities averaged across three outer repeats; the grey diagonal denotes chance discrimination. The shared legend distinguishes the four multivariable models, the two parsimonious logistic benchmarks, and the chance line; reporting-RF AUROCs are in Table 2, and all-model AUROCs are in Supplementary Tables S3–S4. Differences between model families are descriptive unless a paired comparison is explicitly reported. A, amyloid; LR, logistic regression; N, neurodegeneration; PET, positron-emission tomography; RBF-SVM, radial-basis-function support-vector machine; T, tau.

**Table 2.** Cross-axis internal performance of the common random-forest reporting package.

| Outcome | n (positive) | AUROC (95% CI) | AP | Brier | Sens.<sup>a</sup>, % | Spec.<sup>a</sup>, % |
| --- | --- | --- | --- | --- | --- | --- |
| A | 1,097 (496) | 0.9425<br>(0.9271–0.9562) | 0.9356 | 0.0866 | 90.3 | 86.4 |
| T overall | 929 (195) | 0.9495<br>(0.9325–0.9645) | 0.8304 | 0.0682 | 90.4 | 87.5 |
| T within A+ | 361 (161) | 0.8852<br>(0.8485–0.9163) | 0.8593 | 0.1327 | 89.0 | 60.7 |
| Hippocampal N | 1,303 (248) | 0.7865<br>(0.7553–0.8151) | 0.4289 | 0.1285 | 90.2 | 44.3 |
| Joint A+T+ | 852 (161) | 0.9614<br>(0.9484–0.9726) | 0.8408 | 0.0607 | 90.3 | 88.1 |

*Note:* All performance estimates use held-out outer-fold predictions or decisions; each participant was evaluated by outer-fold models that were not trained on that participant. AUROC, AP, and Brier score were calculated once from participant-level probabilities averaged across the three outer repeats; AP was not calculated separately within each repeat and then averaged. CIs are participant-bootstrap percentile intervals conditional on realised averaged out-of-fold predictions. <sup>a</sup>Threshold metrics are percentages using pooled held-out outer-fold decisions at fold-specific thresholds selected only within the corresponding outer-training data to target 90% sensitivity; the held-out sensitivity therefore need not equal 90%. Within a prediction task, lower Brier scores indicate better overall probabilistic performance, but Brier score reflects both calibration and resolution and depends on outcome prevalence. AP, average precision.

### 3.3 Overall and amyloid-positive tau tasks

Restricting the tau task to A+ participants attenuated RF AUROC from 0.9495 to 0.8852. At fold-internal thresholds targeting 90% sensitivity, held-out specificity was 87.5% for T overall and 60.7% for T within A+, while sensitivity was 90.4% and 89.0%, respectively. The attenuation was seen across all four multivariable model families, whose AUROCs ranged from 0.9458 to 0.9495 for T overall and from 0.8819 to 0.8852 within A+. Because T within A+ was a restricted subset with different prevalence and spectrum, this comparison was not assigned a cross-task *P* value and is not evidence that amyloid alone caused the difference.

The 90%-sensitivity/75%-specificity reference was crossed by point estimate for A, T overall, and joint A+T+, but not for T within A+ or N (Table 3). For every outcome, the lower sensitivity confidence limit was below 90%. Only the A comparison approximated the clinical-practice guideline’s target pathology, and even that comparison did not reproduce its specialty-care intended population.

**Table 3.** Descriptive comparison with a 90%-sensitivity/75%-specificity reference.

| Outcome | Sensitivity, % (95% CI) | Specificity, % (95% CI) | Point estimate crosses 90/75 | Sensitivity lower bound ≥ 90% |
| --- | --- | --- | --- | --- |
| A | 90.3 (87.7–92.6) | 86.4 (83.7–89.0) | Yes | No |
| T overall | 90.4 (86.5–94.2) | 87.5 (85.1–89.7) | Yes<sup>a</sup> | No |
| T within A+ | 89.0 (84.4–93.1) | 60.7 (54.5–66.9) | No<sup>a</sup> | No |
| Hippocampal N | 90.2 (86.4–93.4) | 44.3 (41.6–47.0) | No<sup>a</sup> | No |
| Joint A+T+ | 90.3 (85.7–94.4) | 88.1 (85.8–90.4) | Yes<sup>a</sup> | No |

*Note:* Values use pooled held-out outer-fold decisions. Fold-specific thresholds were selected only within the corresponding outer-training data and then applied to the held-out outer fold; a training target of 90% sensitivity does not require held-out sensitivity to equal 90%. The 2025 clinical-practice threshold applies to blood-biomarker assessment of Alzheimer’s disease pathology in objectively cognitively impaired patients in specialty care. <sup>a</sup>For T, N, and joint A+T+, 90/75 is an exploratory common reference and not a guideline-endorsed benchmark.

Qualitative calibration diagnostics for the four multivariable model families are shown in Figure 2; reporting-RF Brier scores are reported in Table 2, and all-model Brier scores are in Supplementary Tables S3–S4. For the reporting RF, joint calibration-intercept point estimates ranged from -0.045 to 0.081 and slope estimates from 0.936 to 1.066 across outcomes; every conditional 95% interval included the corresponding ideal value of 0 or 1 (Supplementary Table S12).

**Fig. 2.**
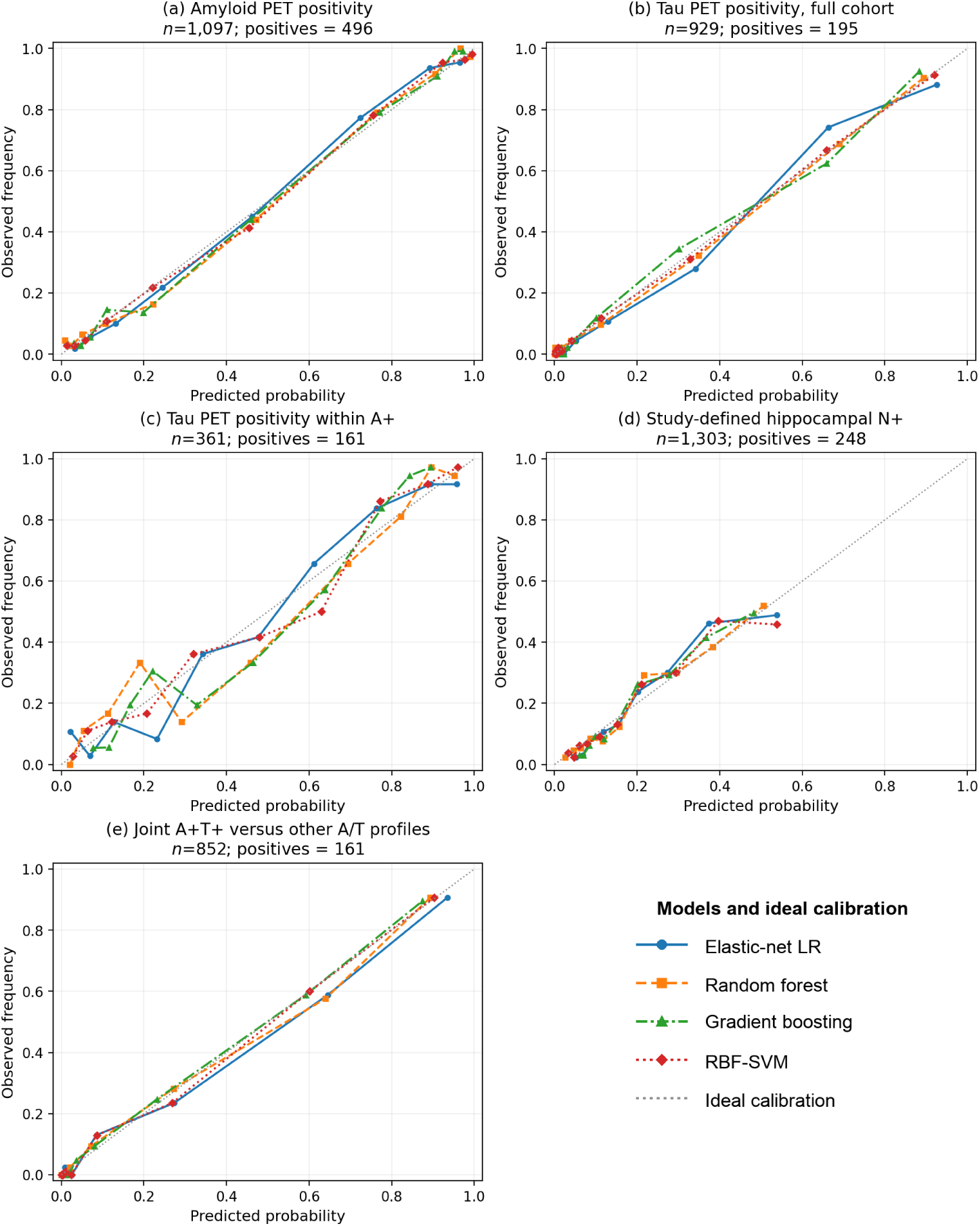
Qualitative internal calibration diagnostics across AT(N) outcomes. Panels show (a) amyloid-PET positivity, (b) tau-PET positivity in the full cohort, (c) tau-PET positivity among amyloid-positive participants, (d) study-defined hippocampal neurodegeneration, and (e) joint A+T+ versus other complete A/T profiles. Curves show equal-frequency decile-binned observed frequencies against participant-level averaged out-of-fold probabilities after fold-internal sigmoid calibration. The grey diagonal denotes ideal agreement, and the shared legend identifies the four multivariable model families. Reporting-RF Brier scores are in Table 2, and all-model Brier scores are in Supplementary Tables S3–S4; they combine calibration and resolution and are not pure calibration measures. Bin-level uncertainty was not estimated; numerical RF calibration diagnostics are in Supplementary Table S12. A, amyloid; LR, logistic regression; N, neurodegeneration; PET, positron-emission tomography; RBF-SVM, radial-basis-function support-vector machine; T, tau.

### 3.4 Predictor-block pattern across axes

Within the ADNI RF analyses, adding plasma A*β*42, A*β*40, and A*β*42/A*β*40 to log(p-tau217) increased AUROC from 0.8942 to 0.9340 for A, 0.9250 to 0.9508 for T overall, 0.8580 to 0.8857 for T within A+, 0.7350 to 0.7659 for N, and 0.9294 to 0.9597 for joint A+T+ (Figure 3). Subsequent addition of demographics and APOE *ε*4 produced smaller changes that varied across prediction tasks. These staged results hold model family constant but remain exploratory predictor-block comparisons; they do not establish independent or causal effects of individual analytes.

**Fig. 3.**
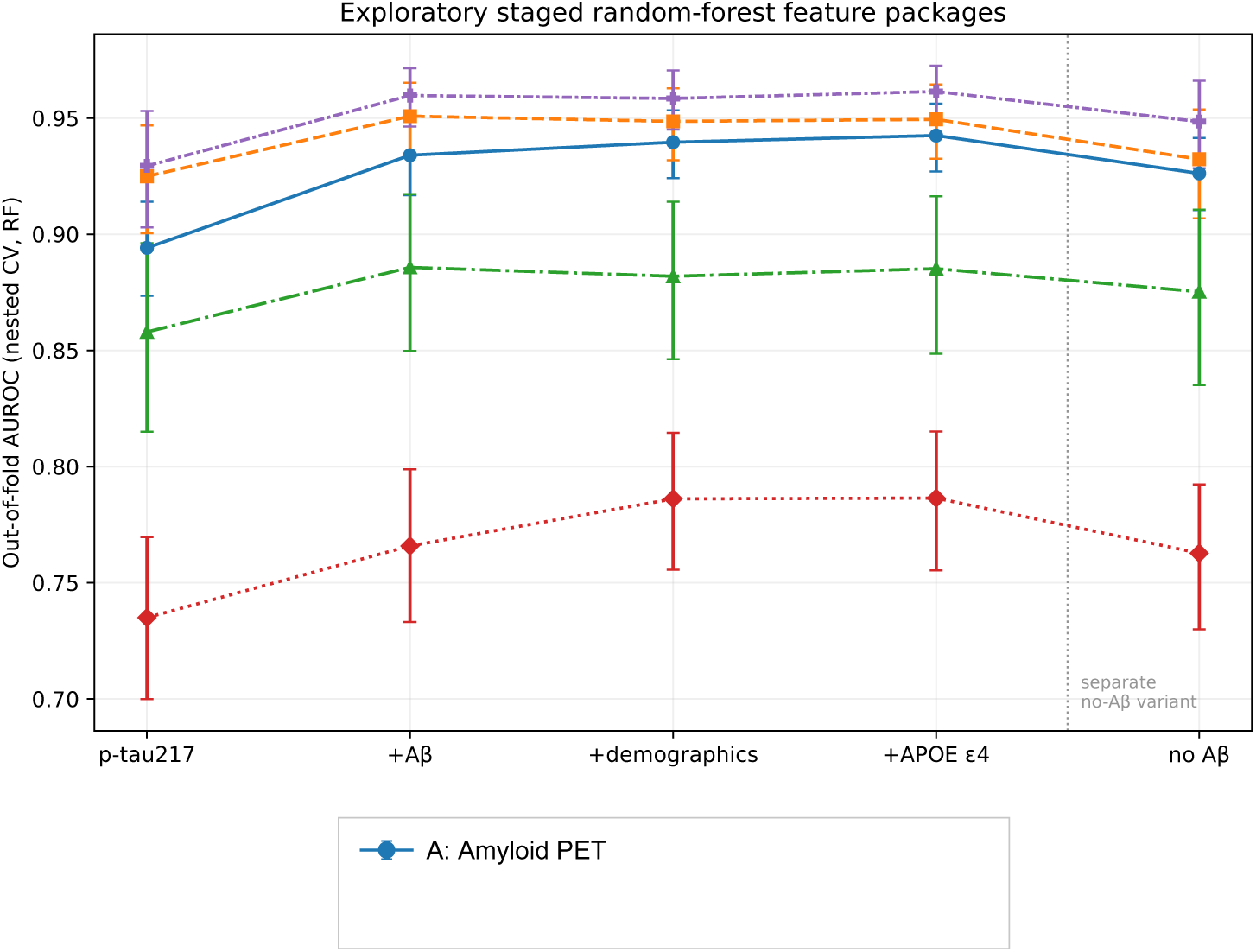
Exploratory staged random-forest feature packages in ADNI. The first four stages are cumulative: p-tau217 alone; addition of plasma A*β*42, A*β*40, and A*β*42/A*β*40; addition of age, sex, and education; and addition of APOE *ε*4. The final point is a separate model without plasma A*β* features. Error bars are participant-bootstrap 95% intervals conditional on fixed out-of-fold predictions. A, amyloid; T, tau; N, study-defined hippocampal neurodegeneration; RF, random forest.

### 3.5 Post-hoc Bio-Hermes A-axis assessment

The common analysis-complete Bio-Hermes cohort included 908 participants; mean age was 72.0 years, 518 (57.0%) were female, and 337 (37.1%) carried at least one APOE *ε*4 allele. Quantified p-tau217 concentrations were available for 743 (81.8%); the other 165 participants retained a censor-aware ordered category (Supplementary Table S13). Of the full cohort, 328 were `AMYCLASV`-positive and 324 were AMYCLAS-positive. Lilly p-tau217 alone achieved an AUROC of 0.894 (95% conditional interval 0.868–0.916) for the primary `AMYCLASV` endpoint. Adding C2N A*β*40 and A*β*42 increased AUROC to 0.916 (0.895–0.935), with paired ΔAUROC 0.022 (0.010–0.035) and ΔBrier -0.011 (-0.017 to -0.005); ΔAP was 0.021 (-0.011 to 0.051). The AMYCLAS sensitivity endpoint was concordant: ΔAUROC 0.022 (0.009–0.034), ΔAP 0.032 (0.002–0.064), and ΔBrier -0.009 (-0.015 to -0.003) (Supplementary Table S11). Adding demographics produced little descriptive change. The APOE-containing stage showed descriptively higher AUROC, but no paired Stage 4-versus-Stage 2 increment was prespecified.

### 3.6 Separate-cohort A4 conditional tau assessment

The formal A4 analysis included 355 official A+ participants with eligible Visit 6 p-tau217 and a complete dated PERSI MTL endpoint; 114 were A4-specific T+ and 241 were T-. Among these 355 participants, 23 p-tau217 results were below the lower limit of quantification, 332 were quantified, and none were above the upper limit of quantification. Blood followed or coincided with the selected tau-PET scan by a median of 12 days (range 0–78). Intervals used 10,000 prespecified participant bootstrap resamples.

Ordinal p-tau217 discriminated A4-specific T status with AUROC 0.7356 (95% participant-bootstrap interval 0.6775–0.7907) and AP 0.5657 (0.4732–0.6677); Spearman correlation with continuous MTL_PERSI_ was 0.4718 (0.3824–0.5528) (Table 4). The 180-day window sensitivity was identical because every observed interval was at most 78 days. Quantified-only and same-day-exclusion sensitivities were fully suppressed under controlled-data disclosure rules because comparison with the primary analysis could reveal protected complementary cells. The strict calendar-day pretreatment subset did not support performance estimation.

**Table 4.** Independent A4 assessment of conditional tau-positivity discrimination.

| Analysis | n (T+) | AUROC (95% interval) | AP (95% interval) | Spearman $\rho$ (95% interval) |
| --- | --- | --- | --- | --- |
| Primary ordinal p-tau217 | 355 (114) | 0.7356<br>(0.6775–0.7907) | 0.5657<br>(0.4732–0.6677) | 0.4718<br>(0.3824–0.5528) |

*Note:* A4-specific T positivity was PERSI MTL SUVR > 1.03. Intervals are 95% participant-bootstrap percentile intervals from 10,000 resamples. AP is average precision calculated with `average_precision_score`. This single-marker A4 assessment is not external validation of the ADNI random-forest package.

### 3.7 A/T profiles and endpoint-definition sensitivity

The three-class A/T elastic-net model had macro one-versus-rest AUROC 0.9107 (0.8935–0.9269), balanced accuracy 0.6833, and log loss 0.5584. For log loss, lower values indicate better probabilistic performance, but the value 0.5584 alone does not establish good calibration without comparison with an appropriate class-proportion benchmark. The four-class configuration contained a sparse A-T+ profile and was retained as descriptive only. Binary shuffled-label AUROCs ranged from 0.476 to 0.533, and the three-class shuffled-label macro AUROC was 0.469. These shuffled-label analyses used one prespecified shuffle per analysis and were negative controls, not formal permutation tests.

As an endpoint-definition diagnostic, we applied alternative labels to existing internal OOF scores without repeating model selection, tuning, calibration, or fitting. Replacing the primary tau definition (SUVR *>* 1.3498184) with the lower SUVR *>* 1.23 stress-test definition changed 202 of 929 T labels, 85 of 361 T-within-A+ labels, and 85 of 852 joint A+T+ labels. Corresponding fixed-score RF AUROCs changed from 0.9495 to 0.7880, from 0.8852 to 0.8016, and from 0.9614 to 0.9238. Replacing residual-based N with the hippocampus/ICV-ratio definition changed 82 of 1,303 labels; RF AUROC changed from 0.7865 to 0.7962. Because the alternative labels define different prediction tasks and estimands, these diagnostics quantify how the existing OOF scores align with those labels; they do not estimate end-to-end performance after retraining (Supplementary Table S8).

### 3.8 Model-family comparisons and design sensitivity analyses

Across the four multivariable model families, A-axis AUROCs ranged from 0.9413 to 0.9450. The RF package had AUROC 0.9425, compared with 0.9118 for log(p-tau217) LR and 0.9303 for log[p-tau217/A*β* 42] LR. The AUROC difference between the RF package and the log(p-tau217) logistic benchmark was 0.0307 (95% unadjusted conditional CI 0.0185–0.0425; Holm-adjusted DeLong *P* = 1.68 *×* 10*^-^*^6^), and the corresponding difference versus the log[p-tau217/A*β*42] logistic benchmark was 0.0122 (0.0022–0.0223; Holm-adjusted *P* = 0.0158). These post-selection package contrasts changed both predictors and model family. Within A+, the RF package showed no evidence of superior discrimination over either parsimonious tau benchmark: ΔAUROC was 0.0041 versus log(p-tau217) LR (95% CI -0.0170 to 0.0271) and -0.0021 versus log[p-tau217/A*β*42] LR (-0.0236 to 0.0192).

A-axis AUROCs ranged from 0.9407 to 0.9444 when the imaging window was changed to *±*180 or *±*730 days or the plasma anchor prioritised completeness. Among 475 cognitively impaired participants, full-cohort OOF predictions had AUROC 0.9593 (0.9406–0.9753), while a separately retrained cognitively impaired-only model had AUROC 0.9524 (0.9330–0.9699). Across RF and elastic-net analyses, log-transform sensitivity ΔAUROCs ranged from -0.00294 to 0.00283. Descriptive subgroup AUROCs ranged from 0.8939 to 0.9592; no formal interaction tests were performed.

## 4 Discussion

This QC-first analysis identified a consistent cross-axis pattern. The plasma-centred RF package strongly discriminated A, overall T, and joint A+T+, whereas performance attenuated for T within A+ and was lower for study-defined hippocampal N. The central finding is therefore the task dependence of plasma prediction across broad pathology detection, conditional tau-positivity discrimination, and downstream structural neurodegeneration, rather than an A-axis model contest.

The contrast between overall and A+-restricted T is consistent with current biomarker biology. Plasma p-tau217 is an early Core 1 marker of AD neuropathologic change rather than a direct measure of insoluble tau-tangle load [3, 11]. Previous studies likewise reported high overall tau-PET discrimination followed by lower performance within A+ samples [5, 6, 8]. Restricting analysis to A+ participants removes the broader amyloid-negative/amyloid-positive contrast and narrows the disease spectrum, creating a stricter conditional tau-positivity task. The observed attenuation is consistent with amyloid-related contribution and spectrum restriction, but does not establish that amyloid caused the high overall T performance.

The A4 assessment provided separate-cohort evidence consistent with the single-marker conditional tau relationship in an A+ cohort. Ordinal Lilly p-tau217 provided moderate discrimination of an A4-specific medial-temporal tau endpoint and showed a concordant positive association with continuous tau burden, consistent with longitudinal A4 evidence linking higher baseline p-tau217 to subsequent tau-PET progression [18]. This evidence was obtained under a different assay representation and endpoint. Because the ADNI multivariable model, preprocessing, calibrated probabilities, and operating thresholds were not transported, A4 was not a validation of the full ADNI RF package. It also lacked an unbiased same-source amyloid-negative comparator and therefore could not reproduce the overall-T versus T-within-A+ contrast. Z-standardising the single A4 score would preserve its ranks and add no discrimination evidence or bridge Lilly and Fujirebio concentration scales.

Strong joint A+T+ discrimination does not conflict with weaker conditional tau-positivity discrimination. The joint task distinguishes a broad pathological profile from other complete A/T profiles and can benefit from strong amyloid-related signal, whereas T within A+ asks whether plasma can further distinguish tau positivity after amyloid positivity is established. The three-class analysis therefore demonstrates descriptive A/T-profile discrimination, not an evaluated enrichment pathway or individual-level replacement of amyloid and tau PET.

The lower performance for hippocampal N is also consistent with the revised framework. Structural N is downstream and nonspecific, and hippocampal volume is influenced by non-Alzheimer pathology, age, and premorbid brain structure [3]. Our endpoint represented one residualised MRI measure rather than the full biological breadth of neurodegeneration. The result does not imply that p-tau217 is unrelated to atrophy; it shows that the plasma-centred package classified this study-defined N endpoint less well than PET-defined A/T outcomes.

In the exploratory ADNI predictor-block analyses, the largest cross-axis change occurred when plasma A*β*42, A*β*40, and their ratio were added to p-tau217; demographics and APOE produced smaller, task-dependent changes. Bio-Hermes provided separate-cohort evidence consistent with modest complementary A-axis information from A*β*40 and A*β*42 beyond p-tau217 under different assay implementations. The pattern was also observed under Bio-Hermes’s primary and alternative PET classifications. However, the Bio-Hermes models were redeveloped internally, and no ADNI probability, calibration model, or threshold was transported. Bio-Hermes therefore supports an A-axis predictor pattern rather than external validation of the ADNI package or its cross-axis performance. Neither analysis isolates independent analyte effects or establishes causality, and p-tau217 implementations differ in numerical scale and performance [29, 30].

The 2025 clinical-practice thresholds require careful scope control because they address blood-based assessment of AD pathology among objectively cognitively impaired patients in specialty care [27]. A was the closest outcome examined here, but ADNI included cognitively unimpaired participants and did not reproduce that intended-care setting; applying the same targets to T, N, and joint A+T+ was therefore exploratory. Point estimates for A, overall T, and joint A+T+ crossed 90% sensitivity and 75% specificity, but no lower sensitivity confidence limit reached 90%. This strict uncertainty criterion supports cautious interpretation: the results do not demonstrate stable threshold attainment, PET/CSF substitution, or clinical readiness.

Tau-related performance also depended on endpoint definition. Relabeling the fixed OOF scores at the lower SUVR 1.23 analysis-code stress-test threshold materially changed T and joint A+T+ AUROCs, consistent with evidence that p-tau217 distinguishes broadly separated biological stages better than adjacent tau stages [7, 8]. Because the original predictions, tuning, and calibration were held fixed, this analysis assessed alignment of the existing score with alternative labels rather than performance after end-to-end redevelopment for the alternative endpoint.

Model-package comparisons were secondary to the cross-axis pattern. On the A axis, the selected RF package had modestly higher AUROC than two parsimonious logistic benchmarks, but predictors and model family changed together; within A+, there was no evidence that RF outperformed either tau benchmark. Repeated nested cross-validation controlled fold-internal tuning leakage but could not address the earlier same-cohort RF selection or the absence of full external evaluation. A future external evaluation should prespecify the target population and endpoints and assess the frozen multivariable model, preprocessing, calibration, and operating rules using a compatible assay implementation.

## 5 Limitations

First, RF selection followed early same-cohort inspection of A-axis results, and the cross-axis synthesis was exploratory rather than a prespecified set of confirmatory analyses. Repeated nested cross-validation controlled fold-internal tuning leakage but could not remove this prior model-family selection history.

Second, comparisons between the full RF package and parsimonious logistic benchmarks changed both predictors and model family, so observed differences cannot be attributed solely to additional biomarkers. The participant bootstrap validly quantified sampling uncertainty conditional on the fixed OOF predictions and labels, but did not propagate uncertainty from model-family selection, reference-threshold estimation, new data partitioning, retuning and refitting, or external transport.

Third, ADNI is a selected research cohort whose clinical spectrum and demographic composition differ from intended-care populations. Outcome-specific eligibility produced different denominators, prevalences, and biomarker spectra, making cross-task contrasts descriptive rather than formal paired comparisons; subgroup analyses also lacked interaction testing. Centre-level clustering and between-centre performance heterogeneity were not modelled. No formal fairness intervention or equity evaluation was undertaken; common preprocessing and QC rules were applied across available demographic groups, but selected-cohort representation and incomplete sociodemo-graphic coverage limit equity and transportability claims. Imaging could occur up to 365 days from the plasma anchor, permitting temporal misclassification despite stable A-axis window sensitivities, and plasma measurements spanned runs and reagent lots without lot-stratified model validation.

Fourth, T and N depended on reference definitions estimated from the same data cut. The external provenance and normalisation equivalence of the lower SUVR 1.23 stress-test threshold were not established, and fixed-score relabeling evaluated label alignment without repeating model development. For N, only MRI records explicitly marked OVERALLQC=Fail were excluded; missing or unrecognised non-Fail QC categories remained eligible, making the MRI QC policy permissive. Age and sex contributed both to the residualised N-label definition and to the predictor package. The resulting performance therefore concerns this operational residual label, not an age- and sex-independent biological construct, and should not be generalised to all forms of neurodegeneration.

Fifth, A4 provided separate-cohort evidence of moderate single-marker conditional tau discrimination under a different assay representation and A4-specific endpoint, but its scope was bounded. Lilly and Fujirebio measurements were not bridged, and the full ADNI model, preprocessing, calibration, probabilities, and thresholds were not transported. The A4 endpoint and 1.03 research cutoff were developed in A4 data; A4 also lacked a suitable same-source amyloid-negative comparator, participant-level confirmation of protocol-scheduled pre-dose timing, and publicly reportable quantified-only and same-day-exclusion sensitivities because of disclosure constraints.

Sixth, Bio-Hermes used internally redeveloped random forests rather than the frozen ADNI package. Its p-tau217 and A*β* assay implementations differed from ADNI, probabilities were uncalibrated, and Stage 1 and Stage 2 estimates were conditional on the common analysis-complete cohort required for all stages. The two PET classifications were alternative definitions within one cohort, not independent external cohorts. The conditional intervals did not propagate full model-development uncertainty. Bio-Hermes did not evaluate T, T within A+, N, or joint A+T+. Thus, Bio-Hermes assessed an A-axis predictor-block pattern and A4 assessed a single-marker conditional-T relationship, but neither evaluated the frozen full ADNI package across AT(N).

Finally, this study did not assess clinical actions, harms, costs, or patient outcomes. Future external evaluation should prespecify the target population and endpoints and test the frozen model and analysis pipeline with a compatible assay implementation in heterogeneous real-world patient populations; prospective clinical-utility studies are separately required before clinical use.

## 6 Conclusions

A plasma-centred multivariable package showed strong internal discrimination for broad PET-defined A, T, and joint A+T+ pathology in ADNI, with lower performance for conditional T discrimination within A+ participants and study-defined hippocampal N. A4 provided separate-cohort evidence consistent with a moderate single-marker conditional tau relationship, while Bio-Hermes provided separate-cohort A-axis evidence that adding A*β*40 and A*β*42 to p-tau217 yielded modest, concordant improvements across two PET classifications. These redeveloped or different-assay assessments do not validate the frozen ADNI package, its calibrated probabilities, or its operating thresholds. Although the ADNI A-axis point estimate crossed the 2025 reference of 90% sensitivity and 75% specificity, its lower sensitivity confidence limit did not reach 90% and the intended specialty-care population was not reproduced. Evaluation of the frozen package in heterogeneous real-world patient populations and prospective assessment of clinical utility remain necessary before clinical thresholds or replacement of PET/MRI can be considered.

## Data Availability

The participant-level data analysed in this study are controlled third-party data and cannot be redistributed by the authors. ADNI data are available to qualified investigators through the LONI Image and Data Archive (https://adni.loni.usc.edu/data-samples/adni-data/) following approval of a data-use application and agreement to the ADNI Data Use Agreement. A4/LEARN data are available to registered investigators through the A4/LEARN data portal (https://www.a4studydata.org/) under the applicable Data Use Agreement. Bio-Hermes-001 data may be requested through the AD Discovery Portal (https://discover.alzheimersdata.org/) and accessed and analysed within AD Workbench subject to the applicable access terms. No participant-level data are included in or redistributed with this article. The analysis code is available from the corresponding author on reasonable request.

https://www.a4studydata.org/

https://discover.alzheimersdata.org/

## List of abbreviations

A: amyloid;
A4: Anti-Amyloid Treatment in Asymptomatic Alzheimer’s Disease;
AD: Alzheimer’s disease;
ADNI: Alzheimer’s Disease Neuroimaging Initiative;
AP: average precision;
AUROC: area under the receiver-operating-characteristic curve;
CDR-SB: Clinical Dementia Rating Sum of Boxes;
CI: confidence interval;
CN: cognitively normal;
ICV: intracranial volume;
LR: logistic regression;
MCI: mild cognitive impairment;
MMSE: Mini-Mental State Examination;
N: neurodegeneration;
OOF: out of fold;
PET: positron-emission tomography;
p-tau217: tau phosphorylated at threonine 217;
RF: random forest;
SUVR: standardized uptake value ratio;
T: tau.

## Additional file

**Additional file 1.** PDF; *Supplementary material for: Performance boundaries of plasma-centred prediction across PET- and MRI-defined AT(N): QC-first internal validation in ADNI with separate axis-specific assessments*. Contains Supplementary Methods, aggregate ADNI cohort and selection summaries, the complete binary model–outcome performance grid, staged analyses, operating points, selected design sensitivities, aggregate multiclass and negative-control results, fixed-score relabeling diagnostics, executed model specifications, the aggregate A4 assessment, the post-hoc Bio-Hermes A-axis assessment and cohort characteristics, ADNI RF calibration diagnostics, and the supplementary precision–recall figure. Participant-level data, private A4 and Bio-Hermes artifacts, and disclosure-sensitive cells are not included. Submission filename: `Additional_file_1.pdf`.

## Declarations

### Ethics approval and consent to participate

This study was a secondary analysis of controlled-access data and involved no new participant contact or data collection. ADNI obtained ethics approval from the institutional review boards of all participating institutions, and all participants or their authorised representatives provided written informed consent in accordance with the Declaration of Helsinki [22, 23]. The A4 study obtained approval from the institutional review board or ethics committee at each participating site; all participants and their study partners provided written informed consent before study initiation, including consent for data collection and sharing [17]. The Bio-Hermes protocol was reviewed and approved by Advarra, a central institutional review board (reference Pro00046018); all participants provided written informed consent, and the study was registered as NCT04733989 [16]. The present analyses were conducted under the respective controlled-data access and data-use agreements.

### Consent for publication

Not applicable.

### Authors’ contributions

JX: Conceptualization, Methodology, Software, Formal analysis, Data curation, Visualization, Writing – original draft, and Writing – review & editing. SM: Supervision, Validation, and Writing – review & editing. FC: Conceptualization, Supervision, Project administration, and Writing – review & editing.

## Acknowledgements

Data collection and sharing for the Alzheimer’s Disease Neuroimaging Initiative (ADNI) is funded by the National Institute on Aging (National Institutes of Health Grant U19AG024904). The grantee organization is the Northern California Institute for Research and Education. In the past, ADNI has also received funding from the National Institute of Biomedical Imaging and Bioengineering, the Canadian Institutes of Health Research, and private-sector contributions through the Foundation for the National Institutes of Health, including generous contributions from AbbVie; Alzheimer’s Association; Alzheimer’s Drug Discovery Foundation; Araclon Biotech; BioClinica, Inc.; Biogen; Bristol-Myers Squibb Company; CereSpir, Inc.; Cogstate; Eisai Inc.; Elan Pharmaceuticals, Inc.; Eli Lilly and Company; EuroImmun; F. Hoffmann-La Roche Ltd and its affiliated company Genentech, Inc.; Fujirebio; GE Healthcare; IXICO Ltd.; Janssen Alzheimer Immunotherapy Research & Development, LLC; Johnson & Johnson Pharmaceutical Research & Development LLC; Lumosity; Lundbeck; Merck & Co., Inc.; Meso Scale Diagnostics, LLC; NeuroRx Research; Neurotrack Technologies; Novartis Pharmaceuticals Corporation; Pfizer Inc.; Piramal Imaging; Servier; Takeda Pharmaceutical Company; and Transition Therapeutics.

The A4 Study was a secondary prevention trial in preclinical Alzheimer’s disease that aimed to slow cognitive decline associated with brain amyloid accumulation in clinically normal older individuals. It was funded by a public–private–philanthropic partnership, including the National Institutes of Health–National Institute on Aging, Eli Lilly and Company, Alzheimer’s Association, Accelerating Medicines Partnership, GHR Foundation, an anonymous foundation, and additional private donors, with in-kind support from Avid Radiopharmaceuticals, Cogstate, Albert Einstein College of Medicine, and the Foundation for Neurologic Diseases. The companion observational Longitudinal Evaluation of Amyloid Risk and Neurodegeneration (LEARN) Study was funded by the Alzheimer’s Association and GHR Foundation. The A4 and LEARN Studies were led by Dr Reisa Sperling at Brigham and Women’s Hospital, Harvard Medical School, and Dr Paul Aisen at the Alzheimer’s Therapeutic Research Institute (ATRI), University of Southern California; coordinated by ATRI; and made available under the auspices of the Alzheimer’s Clinical Trial Consortium through the Global Research & Imaging Platform. The complete A4 Study Team list is available at https://www.actcinfo.org/wp-content/uploads/2024/07/A4-LEARN_Acknowledgements-Longitudinal-List-Final-for-Website.pdf. We acknowledge the dedication of the A4 and LEARN participants and their study partners.

The authors thank the Global Alzheimer’s Platform Foundation, participants, site investigators, and study staff of Bio-Hermes. Bio-Hermes was sponsored by the Global Alzheimer’s Platform Foundation. Eli Lilly and Company provided quantitative readings of ^18^F-florbetapir PET scans and p-tau217 assays. Direct funding was provided by AbbVie, Alzheimer’s Drug Discovery Foundation, Aural Analytics, Biogen, Cognivue, C2N, Gates Ventures, Linus Health, Merck & Co., Quanterix, RetiSpec, and Roche. The Alzheimer’s Disease Data Initiative, Aitia, EMTherapro, Lilly, SomaLogic, and the University of Gothenburg provided in-kind laboratory testing, devices, or testing support. Bio-Hermes data were accessed and analysed through AD Workbench. These source-study acknowledgements do not imply direct funding of the present secondary analysis.

## Alzheimer’s Disease Neuroimaging Initiative

Data used in preparation of this article were obtained from the Alzheimer’s Disease Neuroimaging Initiative (ADNI) database (https://adni.loni.usc.edu). As such, the investigators within the ADNI contributed to the design and implementation of ADNI and/or provided data but did not participate in the analysis or writing of this report. A complete listing of ADNI investigators can be found at: https://adni.loni.usc.edu/wp-content/uploads/how_to_apply/ADNI_Acknowledgement_List.pdf.

