## Additional file 1: Supplementary material for "Performance boundaries of plasma-centred prediction across PET- and MRI-defined AT(N): QC-first internal validation in ADNI with separate axis-specific assessments"

Jiayuan Xu[1], Sofia Michopoulou[2,3], Fumie Costen[1*],
for the Alzheimer's Disease Neuroimaging Initiative

[1]Department of Electrical and Electronic Engineering, University of Manchester, Manchester, United Kingdom.
[2]Medical Physics, University Hospital Southampton NHS Foundation Trust, Southampton, United Kingdom.
[3]Clinical Experimental Sciences, University of Southampton, Southampton General Hospital, Southampton, United Kingdom.

#### Scope and disclosure boundary

This supplement reports aggregate results from the final ADNI analysis after amyloid- and tau-PET quality-control filtering, a post-hoc Bio-Hermes A-axis predictor-block assessment in a cohort separate from ADNI, and the formal A4 conditional tau assessment. Candidate ADNI amyloid- and flortaucipir tau-PET scans were restricted to `qc_flag`=1 or 2 before nearest-date matching. The A4 analysis used official A+ status, ordinal censor-aware p-tau217, and a dated quantitative PERSI medial-temporal endpoint. The Bio-Hermes analysis used Screening blood measures and baseline IXICO PET classifications with scan quality control recorded as pass. Participant identifiers, participant-level predictions, sparse profile cells, the private A4 audit, private Bio-Hermes artifacts, and other disclosure-sensitive data-derived values are not reported. Unless stated otherwise, ADNI estimates come from repeated nested internal cross-validation, Bio-Hermes estimates from repeated cross-validation of models developed within Bio-Hermes, and A4 estimates from a single-marker participant bootstrap.

### Supplementary methods

#### Cohort construction and endpoints

One ADNI row per participant was anchored at the earliest blood draw with p-tau217, A$\beta$42, and A$\beta$40 available. Imaging was matched by nearest absolute date within ±365 days. Amyloid- and tau-PET QC eligibility was applied to raw candidate scans before matching; tau candidates were also restricted to flortaucipir. The released amyloid classification defined A. T positivity used flortaucipir meta-temporal SUVR greater than the mean plus two standard deviations in the dual-PET-QC-filtered A-negative cognitively normal reference group. Overall T and T within A+ were treated as distinct tasks. N positivity used a bilateral hippocampal-volume residual below the reference mean minus two standard deviations after adjustment for intracranial volume, age, and sex. Joint A+T+ contrasted participants positive on both PET endpoints with all other participants having both labels.

#### Release sources and data cleaning

The 21 June 2026 data cut used `UPENN_PLASMA_FUJIREBIO_QUANTERIX_21Jun2026.csv`, `UCBERKELEY_AMY_6MM_21Jun2026.csv`, `UCBERKELEY_TAU_6MM_21Jun2026.csv`, `UCSFFSX7_21Jun2026.csv`, `APOERES_21Jun2026.csv`, `PTDEMOG_21Jun2026.csv`, `DXSUM_21Jun2026.csv`, `MMSE_21Jun2026.csv`, and `CDR_21Jun2026.csv`. Exactly one file per required release prefix was permitted. Raw participant-identifier tokens had to be canonical positive base-10 integers, and every nonmissing anchor or matching date had to parse strictly; malformed values caused the build to fail rather than being normalised or set missing. Clinical variables came from their source releases rather than ADNIMERGE.

Numeric codes $-1$ and $-4$ were set missing for plasma analytes, demographic and clinical variables, and released amyloid status. Plasma A$\beta$42 values above 200 pg/mL and A$\beta$40 values above 3000 pg/mL were set missing; the boundary values were retained. These analyst-defined plausibility bounds were designed to exclude apparent placeholder or entry artifacts and were not interpreted as assay quantification limits. No additional executable concentration bound was applied to p-tau217; all selected p-tau217 values were finite and strictly positive before natural-log transformation. The plasma A$\beta$42/A$\beta$40 ratio was calculated from the cleaned positive concentrations. The earliest dated row with complete p-tau217, A$\beta$42, and A$\beta$40 was selected for each participant. Same-day duplicate release records were collapsed deterministically; conflicting analyte values on the same day caused the build to fail.

Amyloid PET, tau PET, MRI, diagnosis, MMSE, and CDR-SB were matched independently by nearest date within the inclusive $\pm$365-day window. Exact equidistant PET ties selected the earlier scan. Candidate records were not prefiltered on endpoint completeness, and matching did not fall back to a farther record when the selected record lacked the endpoint value. Amyloid candidates were restricted to `qc_flag`=1 or 2 before matching, with flag semantics taken from the controlled release document

ADNI_UCBerkeley_AmyloidPET_Methods_v3_2025-06-30.pdf; the primary A label copied released AMYLOID_STATUS. Tau candidates were restricted first to flortaucipir (FTP) and qc_flag=1 or 2. Within each PET modality, same-day exact scientific replicates were collapsed and conflicting same-day candidates caused the build to fail. Selected amyloid and tau QC flags were retained as provenance and forbidden as predictors.

The primary T cutoff was the dual-PET-QC-filtered A−, cognitively normal reference mean plus two sample standard deviations; positivity required SUVR strictly greater than 1.3498184. The lower fixed SUVR 1.23 definition was retained only as an analysis-code stress test. Its external provenance and normalization equivalence were not established, so it is not presented as a validated literature or clinical cutoff.

For FreeSurfer MRI, rows marked OVERALLQC=Fail were removed; other categories, including missing or unrecognised non-Fail values, remained eligible. Same-day duplicates prioritised complete bilateral hippocampal values, followed by Pass, Partial, Hippocampus Only, and other or missing QC values. Intracranial volume outside 1.0–2.3 million $mm^3$ or either unilateral hippocampal volume outside 1000–6000 $mm^3$ caused all three MRI measurements to be set missing. Sex and education used the first chronological nonmissing value; age was derived from the first parseable birth month and year and the plasma date. APOE $\varepsilon 4$ count was derived from the first nonmissing genotype record.

#### Models and validation

The full predictor package comprised log(p-tau217), A$\beta$42, A$\beta$40, A$\beta$42/A$\beta$40, age, sex, education, and APOE $\varepsilon 4$ count. No predictor values were missing in the final outcome-specific cohorts; every model nevertheless retained fold-internal median imputation and standardisation as a leakage-safe pipeline. Tuning, sigmoid calibration, and threshold selection were also restricted to training data. Evaluation used five outer folds repeated three times and three-fold inner cross-validation. Participant-level held-out probabilities were averaged across repeats for AUROC, average precision (AP), and Brier score. The reporting-RF calibration diagnostic used the same averaged probabilities and an unpenalised joint logistic model of outcome on their log odds; ideal intercept and slope values are 0 and 1. Percentile intervals used 2,000 participant resamples conditional on the realised averaged out-of-fold predictions. These intervals therefore do not account for uncertainty from model-family selection, reference-threshold estimation, or repetition of the full model-development process.

#### Multiplicity and interpretation

Holm adjustment applied only to the two named RF-versus-logistic DeLong tests for the A-axis package comparison. Other comparisons are exploratory and unadjusted. The RF-versus-logistic contrasts change both predictors and model family and therefore compare packages. Staged RF analyses hold model family constant and give exploratory evidence about predictor blocks, not causal effects of individual biomarkers.

#### Bio-Hermes A-axis assessment

Bio-Hermes eligibility required Screening Lilly p-tau217 and C2N A$\beta$40/A$\beta$42, age, sex, education, APOE $\varepsilon$4 allele count, and baseline IXICO `AMYCLASV`, `AMYCLAS`, and `SQC=PASS` records. One common analysis-complete cohort was used for all stages and both outcomes. The primary endpoint was the released IXICO florbetapir visual-read classification `AMYCLASV`; the alternative released IXICO classification `AMYCLAS` was evaluated as an endpoint-definition sensitivity. Both labels were used unchanged. In the parent study, PET scans were centrally read by trained IXICO experts following the manufacturer's read process with access to SUVR values. Disclosure-safe cohort summaries used the same analytic records; Centiloid was descriptive and was not required for eligibility.

Bio-Hermes blood biomarkers were measured in plasma samples; the released predictors comprised Lilly Clinical Diagnostics Laboratory p-tau217 and C2N PrecivityAD A$\beta$40 and A$\beta$42 measurements, analysed on their released scales without cross-assay conversion. The four cumulative random-forest stages used p-tau217 alone; added A$\beta$40 and A$\beta$42; added age, sex, and education; and added APOE $\varepsilon$4 allele count. Within each training fold, assay-censored p-tau217 values were represented as ordered tied categories without numerical substitution, and quantified values retained their numerical order; A$\beta$40 and A$\beta$42 were natural-log transformed. Random forests used 500 trees, balanced-subsample class weights, all available features at each split, one worker, and a three-fold inner grid over minimum leaf sizes 2, 5, and 10. Repeated stratified five-fold cross-validation used three repeats and seed 20261005; the same outer splits were reused across stages within each outcome.

Bio-Hermes models were developed and evaluated entirely within Bio-Hermes using native assay scales and raw random-forest probabilities. AUROC, AP, and Brier score were calculated once per complete OOF repeat and averaged across repeats. Stage 2-minus-Stage 1 was specified within the locked post-hoc assessment before formal fitting. Paired 95% percentile intervals were obtained from 2,000 participant resamples of the fixed repeated-crossfit predictions; models were not refitted during bootstrap.

#### A4 conditional tau assessment

The A4 analysis protocol was locked after exploratory diagnostics but before the first formal analysis run. Eligibility required A4 substudy membership, an official positive screening amyloid-PET status, an eligible Visit 6 protocol-baseline Lilly p-tau217 result, and a complete dated quantitative baseline flortaucipir endpoint. Visit 6 blood was protocol-scheduled before dosing; the release did not provide participant-level blood-draw and infusion clock times. Collection after the treatment-start calendar day was excluded, while collection on that day remained eligible.

Before eligibility filtering, exact duplicate plasma records were collapsed and discordant Visit 6 duplicates caused the analysis to fail. Rescreen identifiers were evaluated as transitive connected components so that one natural person could not enter more than once. Assay results below the lower limit of quantification were tied below all quantified values; quantified values retained their numerical order and ties; values above the upper limit of quantification would have been tied above quantified values. Missing, not-done, invalid, nonpositive, nonfinite, and unknown-status results were excluded. No numeric concentration was assigned to a censored result, and the Lilly measurements were not converted or z-mapped to the ADNI Fujirebio scale.

Quantitative tau-PET candidates had to be baseline visit 4, flortaucipir, affirmatively analysed, and dated. The first eligible baseline scan was selected by actual scan date, with stable scan-number tie handling, before the 365-day blood–scan window was applied. Each scan required one finite PERSI value for each of `entorhinal_lh_VOI`, `entorhinal_rh_VOI`, `parahippocampal_lh_VOI`, and `parahippocampal_rh_VOI`; mixed or conflicting scan metadata or region values caused failure. The composite was

$$\mathrm{MTL}_{\mathrm{PERSI}} = \frac{16146E_L + 15594E_R + 20846P_L + 19991P_R}{72577},$$

and A4-specific T positivity required $\mathrm{MTL}_{\mathrm{PERSI}} > 1.03$. Exact equality was T−. The selected scan had to satisfy $0 \leq$ blood day minus scan day $\leq 365$.

Primary AUROC, AP, and Spearman correlation used the same evaluable participants. AP was calculated with `average_precision_score`. Percentile intervals came from 10,000 participant resamples with seed 20260920; at least 95% of resamples had to yield valid estimates. Prespecified sensitivities retained quantified assay results only, excluded same-day PET–blood pairs, or restricted the imaging window to 180 days. A strict calendar-day pretreatment feasibility subset and threshold equality were retained only in a private aggregate audit. The public evidence required at least 10 participants in each primary T class. A sensitivity row was fully suppressed when any displayed field, or subtraction from another reported total, could recover a nonzero cell below 10.

### Supplementary results

**Table S1** Aggregate cohort construction and reference definitions

| Item | Aggregate result |
|---|---|
| Raw amyloid-PET scans assessed | 4,728 |
| Eligible scans (QC flag 1 or 2) | 4,347 |
| Excluded scans (other, missing, or unassessed flags) | 381 |
| Eligible amyloid scans matched within ±365 days | 1,220 |
| Eligible flortaucipir scans matched within ±365 days after tau QC | 963 |
| Anchored ADNI participants | 1,592 |
| Participants with complete model predictors | 1,522 |
| Amyloid analysis cohort | 1,097 (496 positive) |
| Tau analysis cohort | 929 (195 positive) |
| Tau within amyloid-positive analysis cohort | 361 (161 positive) |
| Study-defined neurodegeneration analysis cohort | 1,303 (248 positive) |
| Joint A+T+ analysis cohort | 852 (161 positive) |
| T reference definition | n=361; cutoff 1.3498184 SUVR |
| N reference definition | n=411; residual cutoff $-1283.6734$ $mm^3$ |
| N ratio sensitivity cutoff | 0.00426101 |

*Note:* Scan-level counts and participant-level counts have different units. Complete-predictor eligibility precedes outcome-specific availability. QC was applied before date matching. This is a partial aggregate flow: the released scan-level counts and final matched participant counts are reported, but intermediate tau-QC attrition is not separately tabulated by participant.

**Table S2** Characteristics by A-axis analysis inclusion

| Characteristic | Included (n=1,097) | Excluded (n=495) |
|---|---|---|
| Age, years, mean (SD) | 72.8 (8.3) | 73.8 (8.3) |
| Female, n (%) | 600 (54.7) | 236 (47.7) |
| Education, years, mean (SD) | 16.4 (2.5) | 16.0 (2.6) |
| APOE $\varepsilon$4 carrier, n/N available (%) | 459/1,097 (41.8) | 205/425 (48.2) |
| Plasma p-tau217, pg/mL, median [IQR] | 0.153 [0.096–0.342] | 0.181 [0.106–0.404] |
| Plasma A$\beta$42, pg/mL, median [IQR] | 26.32 [22.48–29.98] | 25.23 [21.36–29.83] |
| Plasma A$\beta$40, pg/mL, median [IQR] | 300.42 [267.31–336.18] | 297.56 [259.76–338.51] |

*Note:* Included participants met all eligibility requirements for the principal amyloid analysis: the required core model variables and a QC-accepted amyloid-PET scan with an available released amyloid-status label. Excluded participants belonged to the plasma-anchored cohort but did not meet at least one of these requirements. APOE $\varepsilon$4 carrier percentages were calculated among participants with available genotype data. Group characteristics are descriptive; no inferential comparisons were performed.

**Table S3** Internal performance for amyloid and full-cohort tau outcomes

| Outcome | Model | n (positive) | AUROC (95% bootstrap interval) | AP | Brier score |
|---|---|---|---|---|---|
| A | Elastic-net LR | 1,097 (496) | 0.9413 (0.9260–0.9548) | 0.9295 | 0.0891 |
| A | Random forest | 1,097 (496) | 0.9425 (0.9271–0.9562) | 0.9356 | 0.0866 |
| A | Gradient boosting | 1,097 (496) | 0.9440 (0.9293–0.9570) | 0.9422 | 0.0870 |
| A | RBF-SVM | 1,097 (496) | 0.9450 (0.9306–0.9579) | 0.9402 | 0.0864 |
| A | log(p-tau217) LR | 1,097 (496) | 0.9118 (0.8931–0.9294) | 0.8897 | 0.1152 |
| A | log[p-tau217/A$\beta$42] LR | 1,097 (496) | 0.9303 (0.9131–0.9460) | 0.9023 | 0.0987 |
| T | Elastic-net LR | 929 (195) | 0.9458 (0.9268–0.9625) | 0.8140 | 0.0678 |
| T | Random forest | 929 (195) | 0.9495 (0.9325–0.9645) | 0.8304 | 0.0682 |
| T | Gradient boosting | 929 (195) | 0.9460 (0.9266–0.9628) | 0.8397 | 0.0695 |
| T | RBF-SVM | 929 (195) | 0.9492 (0.9325–0.9643) | 0.8489 | 0.0679 |
| T | log(p-tau217) LR | 929 (195) | 0.9424 (0.9231–0.9602) | 0.7974 | 0.0686 |
| T | log[p-tau217/A$\beta$42] LR | 929 (195) | 0.9495 (0.9333–0.9645) | 0.8134 | 0.0678 |

*Note:* Intervals are 95% conditional participant-bootstrap percentile intervals. A, amyloid; T, tau; AP, average precision; LR, logistic regression; RBF-SVM, radial-basis-function support-vector machine.

**Table S4** Internal performance for the remaining binary outcomes

| Outcome | Model | n (positive) | AUROC (95% bootstrap interval) | AP | Brier score |
|---|---|---|---|---|---|
| T within A+ | Elastic-net LR | 361 (161) | 0.8835 (0.8460–0.9179) | 0.8466 | 0.1332 |
| T within A+ | Random forest | 361 (161) | 0.8852 (0.8485–0.9163) | 0.8593 | 0.1327 |
| T within A+ | Gradient boosting | 361 (161) | 0.8819 (0.8444–0.9131) | 0.8615 | 0.1363 |
| T within A+ | RBF-SVM | 361 (161) | 0.8826 (0.8470–0.9151) | 0.8715 | 0.1365 |
| T within A+ | log(p-tau217) LR | 361 (161) | 0.8811 (0.8421–0.9163) | 0.8345 | 0.1334 |
| T within A+ | log[p-tau217/A$\beta$42] LR | 361 (161) | 0.8873 (0.8524–0.9206) | 0.8705 | 0.1313 |
| N | Elastic-net LR | 1,303 (248) | 0.7911 (0.7611–0.8190) | 0.4304 | 0.1289 |
| N | Random forest | 1,303 (248) | 0.7865 (0.7553–0.8151) | 0.4289 | 0.1285 |
| N | Gradient boosting | 1,303 (248) | 0.7780 (0.7469–0.8064) | 0.4303 | 0.1305 |
| N | RBF-SVM | 1,303 (248) | 0.7836 (0.7528–0.8127) | 0.4235 | 0.1294 |
| N | log(p-tau217) LR | 1,303 (248) | 0.7655 (0.7310–0.7983) | 0.4257 | 0.1311 |
| N | log[p-tau217/A$\beta$42] LR | 1,303 (248) | 0.7671 (0.7334–0.7994) | 0.4177 | 0.1319 |
| Joint A+T+ | Elastic-net LR | 852 (161) | 0.9577 (0.9438–0.9702) | 0.8437 | 0.0627 |
| Joint A+T+ | Random forest | 852 (161) | 0.9614 (0.9484–0.9726) | 0.8408 | 0.0607 |
| Joint A+T+ | Gradient boosting | 852 (161) | 0.9567 (0.9422–0.9692) | 0.8426 | 0.0633 |
| Joint A+T+ | RBF-SVM | 852 (161) | 0.9585 (0.9448–0.9707) | 0.8480 | 0.0626 |
| Joint A+T+ | log(p-tau217) LR | 852 (161) | 0.9495 (0.9309–0.9670) | 0.7918 | 0.0632 |
| Joint A+T+ | log[p-tau217/A$\beta$42] LR | 852 (161) | 0.9564 (0.9419–0.9698) | 0.8042 | 0.0626 |

*Note:* N is the study-defined hippocampal atrophy outcome. Intervals are conditional participant-bootstrap percentile intervals. AP, average precision.

**Table S5** Exploratory staged random-forest analyses

| Outcome | Stage 1: log(p-tau217) | Stage 2: + plasma A$\beta$ | Stage 3: + demographics | Stage 4: + APOE $\varepsilon$4 | Full package without A$\beta$ |
|---|---|---|---|---|---|
| A | 0.8942 | 0.9340 | 0.9396 | 0.9425 | 0.9262 |
| T | 0.9250 | 0.9508 | 0.9486 | 0.9495 | 0.9323 |
| T within A+ | 0.8580 | 0.8857 | 0.8819 | 0.8852 | 0.8753 |
| N | 0.7350 | 0.7659 | 0.7861 | 0.7865 | 0.7627 |
| Joint A+T+ | 0.9294 | 0.9597 | 0.9585 | 0.9614 | 0.9485 |

*Note:* Cells are rounded AUROCs. Stages 1–4 are cumulative predictor packages. For A, adjacent unrounded $\Delta$AUROCs were +0.0399 (95% interval 0.0269–0.0534), +0.0056 (0.0015–0.0096), and +0.0029 (−0.0008 to 0.0069). All staged comparisons are exploratory and unadjusted.

**Table S6** A-axis operating points and design sensitivities

| Analysis | Aggregate result |
|---|---|
| RF Youden operating point | Median fold threshold 0.4616; sensitivity 86.3%; specificity 89.7%; PPV 87.4%; NPV 88.8%; accuracy 88.2% |
| RF target $\geq$ 90% sensitivity | Median fold threshold 0.3661 (fold IQR 0.3360–0.3844); sensitivity 90.3% (95% interval 87.7–92.6%); specificity 86.4% (83.7–89.0%); PPV 84.6%; NPV 91.5% |
| Two-cutoff 90% sensitivity/90% specificity scenario | 51.75% rule-out, 4.13% indeterminate, 44.12% rule-in; rule-out NPV 91.5%; rule-in PPV 88.0% |
| Two-cutoff 95% sensitivity/95% specificity scenario | 39.26% rule-out, 22.24% indeterminate, 38.50% rule-in; rule-out NPV 94.0%; rule-in PPV 92.7% |
| ±180-day earliest anchor | AUROC 0.9407 (0.9250–0.9558) |
| ±730-day earliest anchor | AUROC 0.9442 (0.9305–0.9564) |
| ±365-day most-complete anchor | AUROC 0.9444 (0.9300–0.9584) |
| Cognitively impaired, full-cohort OOF scores | n=475 (304 positive); AUROC 0.9593 (0.9406–0.9753) |
| Cognitively impaired, nested retraining | n=475 (304 positive); AUROC 0.9524 (0.9330–0.9699) |

*Note:* Thresholds were selected inside training folds. Two-cutoff percentages are hypothetical internal referral scenarios rather than observed clinical savings. AUROC, area under the receiver-operating-characteristic curve; IQR, interquartile range; OOF, out of fold; PPV, positive predictive value; NPV, negative predictive value; RF, random forest.

**Table S7** Exploratory multiclass and negative-control analyses

| Analysis | Aggregate result |
|---|---|
| Three-class A/T elastic-net model | Macro one-versus-rest AUROC 0.9107 (0.8935–0.9269); balanced accuracy 0.6833; log loss 0.5584 |
| Binary shuffled-label controls | AUROCs 0.476–0.533 across the five outcomes |
| Three-class shuffled-label control | Macro one-versus-rest AUROC 0.469 |

*Note:* To protect participant confidentiality and avoid unstable inference, four-class profile metrics, sparse counts, and complementary confusion cells are suppressed. Multiclass results are descriptive and were not used for profile-level clinical claims.

**Table S8** Fixed-score alternative-label diagnostics for the reporting random forest

| Outcome | n | Positive, primary → alternative | Labels changed | RF AUROC, primary → alternative |
|---|---|---|---|---|
| Full-cohort T | 929 | 195 → 397 | 202 | 0.9495 → 0.7880 |
| T within A+ | 361 | 161 → 246 | 85 | 0.8852 → 0.8016 |
| Joint A+T+ | 852 | 161 → 246 | 85 | 0.9614 → 0.9238 |
| Study-defined N | 1,303 | 248 → 256 | 82 | 0.7865 → 0.7962 |

*Note:* For T-related outcomes, the primary reference-derived cutoff (SUVR 1.3498184) was replaced by the lower SUVR $>$ 1.23 analysis-code stress test; external provenance and normalization equivalence for 1.23 were not established. For N, the residual definition (cutoff $-1283.6734$ mm$^3$) was replaced by the hippocampus/ICV-ratio definition (cutoff 0.00426101). Existing internal out-of-fold scores were held fixed while labels were changed. Models were not reselected, retuned, recalibrated, or retrained; these are endpoint-definition diagnostics, not end-to-end nested-cross-validation sensitivity estimates. Change totals overlap across T-related outcomes and must not be summed.

**Table S9**: Executed model and validation specifications

| Component | Executed specification |
|---|---|
| Software | Python 3.9.6; NumPy 2.0.2; pandas 2.3.3; SciPy 1.13.1; scikit-learn 1.6.1; joblib 1.5.3. |
| Predictors and preprocessing | Full models used log(p-tau217), A$\beta$42, A$\beta$40, A$\beta$42/A$\beta$40, age, sex (female=1), education, and APOE $\varepsilon$4 allele count. Every estimator used a pipeline with training-fold median imputation followed by standardisation; this included tree models and binary/count predictors. |
| Elastic-net logistic regression | Custom binary single-logit split-L-BFGS-B implementation; elastic-net penalty; unpenalised intercept; balanced class weights; random state 42; maximum 50,000 iterations; tolerance $10^{-8}$. Grid: $C \in \{10^{-4}, 10^{-2}, 1, 10^{2}, 10^{4}\}$ and `l1_ratio` $\in \{0.1, 0.5, 0.9\}$ (15 candidates). |
| Random forest | 500 trees; balanced-subsample class weights; random state 42; one worker. Grid: minimum leaf size $\in \{2, 5, 10\}$ and maximum features $\in \{\text{sqrt}, 0.5\}$ (6 candidates); other scikit-learn 1.6.1 defaults were retained. |
| Gradient boosting | 300 estimators; subsample 0.8; no class weighting; random state 42. Grid: learning rate $\in \{0.03, 0.05, 0.1\}$ and maximum depth $\in \{2, 3\}$ (6 candidates); other scikit-learn 1.6.1 defaults were retained. |
| RBF support-vector machine | RBF kernel; balanced class weights; internal probability fitting disabled; random state 42. Grid: $C \in \{0.1, 1, 10\}$ and $\gamma \in \{\text{scale}, 0.03, 0.1\}$ (9 candidates). The decision function fed the separate sigmoid calibrator. |
| Parsimonious benchmarks | Separate scikit-learn L2-logistic models used either log(p-tau217) or log[p-tau217/A$\beta$42] alone; $C = 1$, `lbfgs`, balanced class weights, maximum 2,000 iterations, tolerance $10^{-4}$, and random state 42. |

Continued on next page

Table S9 continued

| Component | Executed specification |
|---|---|
| Hyperparameter selection | Within each outer training set, three-fold stratified inner cross-validation used ROC-AUC, shuffling, random state $42 + k$ for $k = 0, \ldots, 14$ indexing the 15 outer folds across repeats, one worker, and no automatic refit. The selected parameters were applied to fresh clones. Exact ties selected the first candidate in deterministic parameter-grid order; candidate-level score margins were not retained. |
| Calibration | For each model and outer fold, a separate three-fold stratified split of the outer-training set used random state $1042 + k$ and the selected hyperparameters to generate raw out-of-fold scores. Elastic-net and both benchmark logistic models and the RBF-SVM supplied decision-function scores; random-forest and gradient-boosting probabilities were clipped to $[10^{-6}, 1 - 10^{-6}]$ and converted to log odds. An untuned L2-logistic sigmoid calibrator used $C = 1$, `lbfgs`, tolerance $10^{-4}$, no class weighting, maximum 1,000 iterations, and random state 42. It was fitted to all inner out-of-fold scores and labels. The selected base pipeline was refitted on the complete outer-training set, and the fixed calibrator transformed its outer-test scores. |
| Threshold selection | Youden $J = \text{sensitivity} + \text{specificity} - 1$, sensitivity targets 0.90 and 0.95, and specificity targets 0.90 and 0.95 were selected from calibrated inner out-of-fold probabilities using the descending thresholds returned by scikit-learn's ROC curve. Youden selected the highest-threshold maximiser; sensitivity selected the highest threshold meeting the target (or the lowest available threshold if none did), and specificity selected the lowest threshold meeting the target (or the highest available threshold if none did). Outer-test decisions used probability greater than or equal to the fold-specific threshold. |
| Outer evaluation and aggregation | Label 1 was the positive class and larger scores denoted greater predicted positivity. Repeated stratified five-fold cross-validation with three repeats and random state 42 produced 15 outer test folds. Each participant received one held-out probability and decision per repeat. The three probabilities were averaged before calculating AUROC, AP, Brier score, and paired model comparisons. Threshold metrics were calculated from one confusion matrix over the pooled $3n$ repeat-specific decisions, so each participant contributed three equally weighted decisions. |
| Uncertainty and multiplicity | AUROC intervals used 2,000 ordinary participant resamples with replacement from fixed averaged out-of-fold scores; one-class resamples were skipped, and the 2.5th and 97.5th percentiles formed 95% intervals. Paired ΔAUROC intervals used 2,000 class-stratified paired-participant resamples with replacement. Two-sided paired DeLong tests treated the averaged scores as fixed and did not account for model selection or overlapping training sets. Sensitivity and specificity intervals used 2,000 participant-cluster resamples carrying all three decisions. Only the two named amyloid RF-versus-benchmark DeLong $P$ values received Holm correction. AP and Brier score had no formal bootstrap intervals. |

**Table S10** Aggregate A4 conditional tau assessment

| Item | Aggregate result |
|---|---|
| A4 plasma-release participants | 1,127 |
| Visit 6 participants | 1,116 |
| Eligible protocol-baseline plasma | 1,080 |
| Official amyloid-positive status | 1,080 |
| Complete dated PERSI MTL endpoint | 355 |
| A4-specific T outcome | 114 T+; 241 T− |
| Ordinal assay status | 23 below LLOQ; 332 quantified; 0 above ULOQ |
| Blood minus selected tau-PET interval | n=355; median 12 days; range 0–78 |
| Blood minus treatment-start interval | n=355; median 0 days; range −7 to 0 |
| Primary ordinal p-tau217 | AUROC 0.7356 (95% interval 0.6775–0.7907); AP 0.5657 (0.4732–0.6677); Spearman $\rho = 0.4718$ (0.3824–0.5528) |
| Bootstrap completion | 10,000/10,000 valid participant resamples |
| 180-day imaging-window sensitivity | Identical to the primary analysis |
| Quantified-only sensitivity | Fully suppressed because of complementary small-cell disclosure risk |
| Exclude same-day PET–blood sensitivity | Fully suppressed because of complementary small-cell disclosure risk |
| Strict calendar-day pretreatment subset | Performance not estimable; private flow audit only |

*Note:* Positivity required PERSI MTL SUVR $>$ 1.03. AP is average precision calculated with `average_precision_score`. Intervals are participant-bootstrap percentile intervals. The transition from 1,080 eligible A+ plasma participants to 355 complete dated PERSI MTL endpoints bundles imaging availability, region completeness, date, and interval requirements. The treatment-start interval uses calendar days; same-day blood was protocol-scheduled before dosing, but participant-level clock order was unavailable. LLOQ, lower limit of quantification; ULOQ, upper limit of quantification.

**Table S11** Post-hoc within-cohort Bio-Hermes A-axis predictor-block assessment

| Endpoint | Predictor stage or contrast | AUROC (95% interval) | AP (95% interval) | Brier (95% interval) |
|---|---|---|---|---|
| `AMYCLASV` (primary) | Stage 1: p-tau217 | 0.894 (0.868–0.916) | 0.834 (0.787–0.876) | 0.115 (0.100–0.130) |
| | Stage 2: + A$\beta$40, A$\beta$42 | 0.916 (0.895–0.935) | 0.855 (0.814–0.895) | 0.104 (0.091–0.118) |
| | Stage 3: + demographics | 0.915 (0.893–0.934) | 0.843 (0.799–0.888) | 0.104 (0.091–0.118) |
| | Stage 4: + APOE $\varepsilon$4 | 0.924 (0.905–0.941) | 0.858 (0.815–0.903) | 0.101 (0.088–0.114) |
| | Stage 2 − Stage 1 | 0.022 (0.010–0.035) | 0.021 (−0.011–0.051) | −0.011 (−0.017–−0.005) |
| `AMYCLAS` (sensitivity) | Stage 1: p-tau217 | 0.893 (0.869–0.917) | 0.822 (0.775–0.866) | 0.115 (0.100–0.130) |
| | Stage 2: + A$\beta$40, A$\beta$42 | 0.915 (0.893–0.934) | 0.854 (0.810–0.893) | 0.106 (0.092–0.121) |
| | Stage 3: + demographics | 0.911 (0.889–0.931) | 0.842 (0.795–0.885) | 0.106 (0.092–0.121) |
| | Stage 4: + APOE $\varepsilon$4 | 0.923 (0.905–0.940) | 0.852 (0.807–0.896) | 0.103 (0.090–0.117) |
| | Stage 2 − Stage 1 | 0.022 (0.009–0.034) | 0.032 (0.002–0.064) | −0.009 (−0.015–−0.003) |

*Note:* The same common analysis-complete cohort of 908 participants was used for every stage and both endpoints; 328 were `AMYCLASV`-positive and 324 were `AMYCLAS`-positive. Stage metrics are means of complete OOF metrics across three repeats. Intervals are bootstrap percentile intervals from 2,000 paired participant resamples of the fixed repeated-crossfit predictions. Models were developed and evaluated within Bio-Hermes using native assay scales and raw random-forest probabilities. AP, average precision.

**Table S12** Calibration diagnostics for the ADNI reporting random forest

| Outcome | n (positive) | Calibration intercept (95% interval) | Calibration slope (95% interval) |
|---|---|---|---|
| A | 1,097 (496) | 0.010 (−0.174–0.195) | 0.983 (0.876–1.118) |
| T overall | 929 (195) | −0.034 (−0.275–0.224) | 0.975 (0.848–1.145) |
| T within A+ | 361 (161) | −0.025 (−0.302–0.254) | 0.936 (0.785–1.156) |
| Hippocampal N | 1,303 (248) | 0.081 (−0.155–0.320) | 1.066 (0.911–1.241) |
| Joint A+T+ | 852 (161) | −0.045 (−0.306–0.245) | 0.983 (0.841–1.168) |

*Note:* Intercept and slope were estimated jointly by unpenalised logistic calibration of each outcome on the log odds of participant-level probabilities averaged across the three held-out outer repeats. Ideal values are 0 and 1, respectively. Intervals are 95% participant-bootstrap percentile intervals from 2,000 resamples, conditional on the fixed averaged out-of-fold predictions.

**Table S13** Bio-Hermes cohort characteristics and analytic completeness

| Characteristic | Aggregate result |
|---|---|
| Participants | 908 |
| Primary `AMYCLASV` positive | 328 (36.1%) |
| Sensitivity `AMYCLAS` positive | 324 (35.7%) |
| Age, years, mean (SD) | 72.0 (6.7) |
| Female | 518 (57.0%) |
| Education, years, mean (SD) | 15.5 (2.7) |
| APOE $\varepsilon 4$ carrier | 337 (37.1%) |
| Quantified p-tau217 concentration available | 743 (81.8%) |
| Censor-aware category without a quantified p-tau217 concentration | 165 (18.2%) |
| C2N A$\beta$40, median [IQR] | 483.51 [440.98–543.75] |
| C2N A$\beta$42, median [IQR] | 47.05 [41.85–53.13] |
| Centiloid, median [IQR] | 10.9 [−5.0–55.9] (n=886) |

*Note:* All model variables and both binary IXICO PET classifications were complete in the common 908-participant cohort. Quantified p-tau217 concentration was not required when an assay-censored result could be represented by its ordered category. Centiloid was available for 886 participants and was descriptive only; it was not a model predictor or eligibility requirement. A$\beta$ summaries use the released concentration scales. IQR, interquartile range.

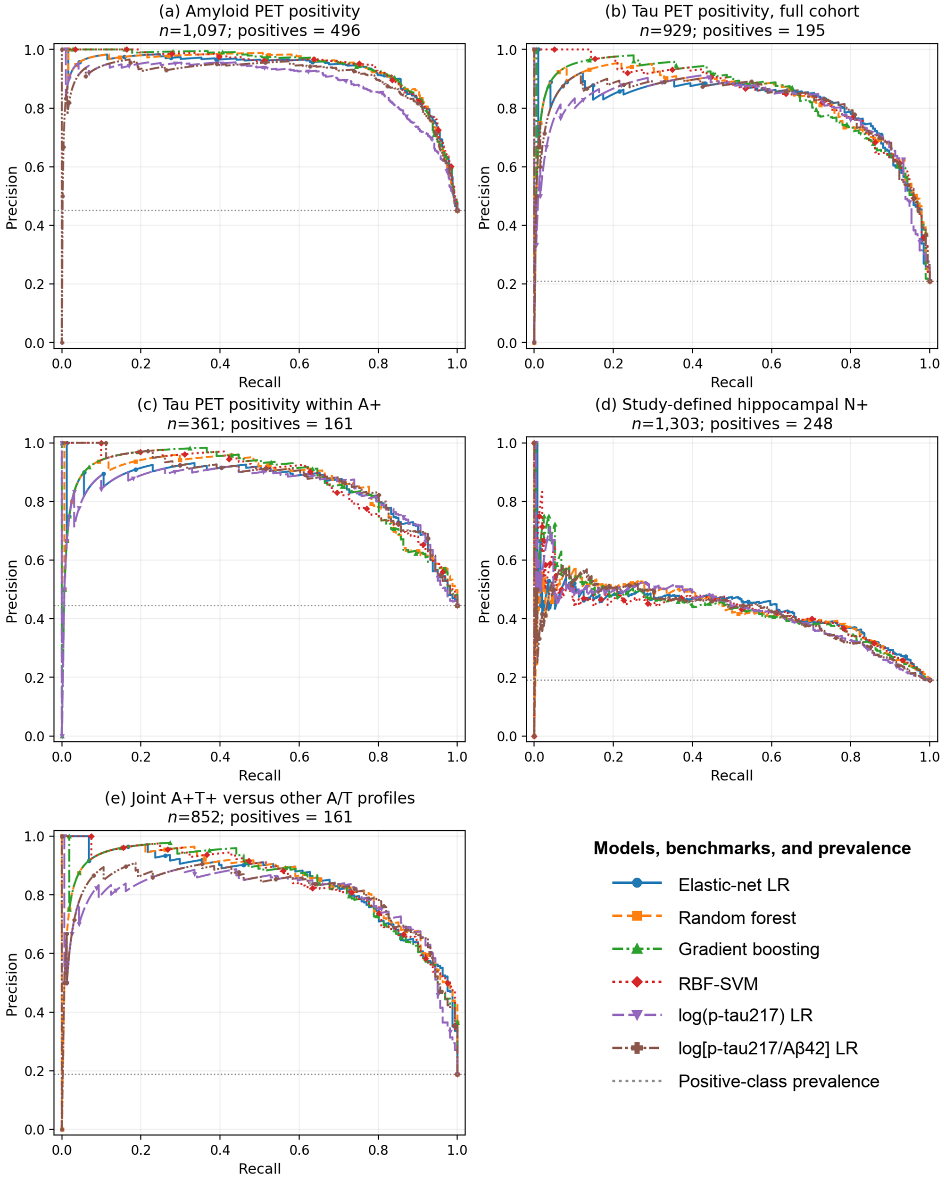


**Fig. S1** Internal-validation precision–recall curves. Panels show (a) amyloid-PET positivity, (b) tau-PET positivity in the full cohort, (c) tau-PET positivity among amyloid-positive participants, (d) study-defined hippocampal neurodegeneration, and (e) joint A+T+ versus other complete A/T profiles; grey dotted horizontal lines denote positive-class prevalence. Curves use participant-level out-of-fold probabilities averaged across three outer repeats. The shared legend distinguishes the four multivariable models, two parsimonious logistic benchmarks, and the positive-class prevalence reference; average precision values are in Supplementary Tables S3 and S4. A, amyloid; LR, logistic regression; N, neurodegeneration; PET, positron-emission tomography; RBF-SVM, radial-basis-function support-vector machine; T, tau.
